# Improved Identification of Large-effect Rare Genetic Variants using Haplotype Aggregated Allele-specific Expression Data

**DOI:** 10.64898/2025.12.16.25341855

**Authors:** Kaushik Ram Ganapathy, Martin Broly, Sarah Silverstein, Marcela Mendoza, Eric Song, Bence Kotis, Paul Hoffman, PCGC Consortium, Ali Torkamani, David R Adams, Carsten Bonnemann, Tuuli Lappalainen, Pejman Mohammadi

## Abstract

Allele-specific expression (ASE) outlier detection is a powerful tool for identifying genes affected by large effect rare genetic regulatory variants but suffers from data sparsity and noisy signal in low-count genes. Genome phasing can be utilized to aggregate ASE signal along haplotypes to alleviate both sparsity and noise. Yet statistical tools for utilizing haplotype-level ASE data for rare variant interpretation are lacking. Here, we present ANEVA-h, to quantify the amount of genetic variation in gene expression from haplotype-level ASE data in a population, enabling more accurate and comprehensive detection of regulatory effects. We apply ANEVA-h to GTEx project data, along with a compatible dosage outlier test, to show an over 2-fold increase in the number of testable genes, reduction of spurious outlier calls, and improved enrichment for rare high-impact variants. In clinical cohorts of neuromuscular and congenital heart disease, it enhances gene prioritization and identifies candidate diagnoses missed by DROP-MAE and ANEVA. Finally, we analyze globally diverse populations to characterize the impact of ancestry background in reference and the test population. We provide tools and data necessary to facilitate integration of haplotype level ASE outlier testing in rare variant interpretation pipelines

## Introduction

The integration of genome and transcriptome sequencing has enabled the systematic identification of functional noncoding variants by linking genetic differences to transcriptional consequences [1–12]. Among these, allele-specific expression (ASE)—the expression associated with each allele at a transcribed heterozygous sites—serves as a direct readout of differences in cis-regulatory activity [5, 13]. Due to its high heritability, ASE data has is an exceptional assay for identifying genes likely affected by large effect rare or de-novo cis-regulatory variants [14]. However, current frameworks for ASE analysis primarily rely on SNP-level ASE measurements, which suffer from key limitations.

SNP-level ASE data often has relatively low counts and is inherently sparse with each variant allowing for generation of ASE data in at most half of the individuals under Hardy-Weinberg equilibrium. Furthermore, depending on where on a gene the aseSNP is located, and its linkage disequilibrium with non-coding regulatory variants that drive allelic imbalance, the observed ASE signal can vary. A common strategy to associate a single ASE readout to a gene is to select the highest-expressed heterozygous SNP as a proxy for the overall regulatory signal. While this maximizes data availability, it discards a large fraction of the data, and it excludes many genes entirely: those lacking a single heterozygous site with sufficient read are dropped from analysis [8]. This results in substantial signal loss especially in lowly expressed genes or genes with few transcribed heterozygous sites [7, 8]. And for retained genes, single-SNP measurements fail to capture distributed regulatory effects across the locus and can yield inconsistent or misleading signals when multiple variants exert opposing cis effects [15]. Technical artifacts such as reference mapping bias and local sequence context further confound interpretation [16]. These limitations are particularly acute in genes with complex structure, long transcripts, or tissue-specific splicing, ultimately reducing the sensitivity, interpretability, and reliability of SNP-level ASE for regulatory variant detection.

Aggregating ASE signal across multiple aseSNPs and along phased haplotypes has been used to mitigate limitations of conventional ASE data but the statistical methods to utilize the haplotype-level ASE data for identifying genes with excessive allelic imbalance are still lacking. To address this, we introduce ANEVA-h, a statistical framework that aggregates ASE signal across phased haplotypes to estimate the expected genetic variance in gene expression in a population enabling more robust regulatory outlier detection, and significantly expanding the number of tested genes in a given sample. We apply ANEVA-h to large-scale population datasets—including the Genotype-Tissue Expression (GTEx) project and the MAGE cohort of globally diverse individuals, and to two clinical cohorts of mendelian muscular dystrophies and myopathies and congenital heart disease [7, 8, 11, 17–19]. Across these settings, we demonstrate that haplotype-level ASE improves sensitivity to regulatory effects, increases the number of testable genes, and enables the detection of large-effect rare variants missed by existing methods.

## Results

### Haplotype Aggregation Improves ASE Signal

Summarizing of ASE signals at the gene level has traditionally relied on selecting the SNP with the highest total expression within each gene [1, 8]. However, this approach discards the ASE signal when available in multiple SNPs distributed across the gene, and can pick regulatory discordant SNPs in different individuals causing elevated false positive rate in certain genes [8].In contrast, haplotype-level aggregation of ASE data offers a more comprehensive strategy by combining ASE signal across phased SNPs within each gene, improving coverage and providing a more stable estimate of allelic imbalance[1, 15]. phASER, a widely used tool for haplotype-based ASE analysis, implements this approach by combining population-based genotype phasing with read-backed phasing, enabling accurate assignment of RNA-seq reads to parental haplotypes. It also corrects for technical artifacts such as double-counting of overlapping reads and ambiguous fragment assignments, ensuring reliable quantification of haplotypic expression [15].

To explore the potential of haplotype aggregated ASE data, we first compared variant and haplotype-aggregatedASE data from the GTEx consortium v8 across 49 tissues generated using ASEReadCounter and phASER, respectively [15, 20]. On average, we identified an additional 1198 genes per sample with total ASE count above 10 in haplotype-level data (Fig 1B). Further, among the shared genes, haplotype aggregation showed increased total ASE counts with increased number of available ASE SNPs (Spearman-rho = 0.82) in-line with those previously reported in Castel et. al. (Fig. 1C, [15]).

**Figure 1.**
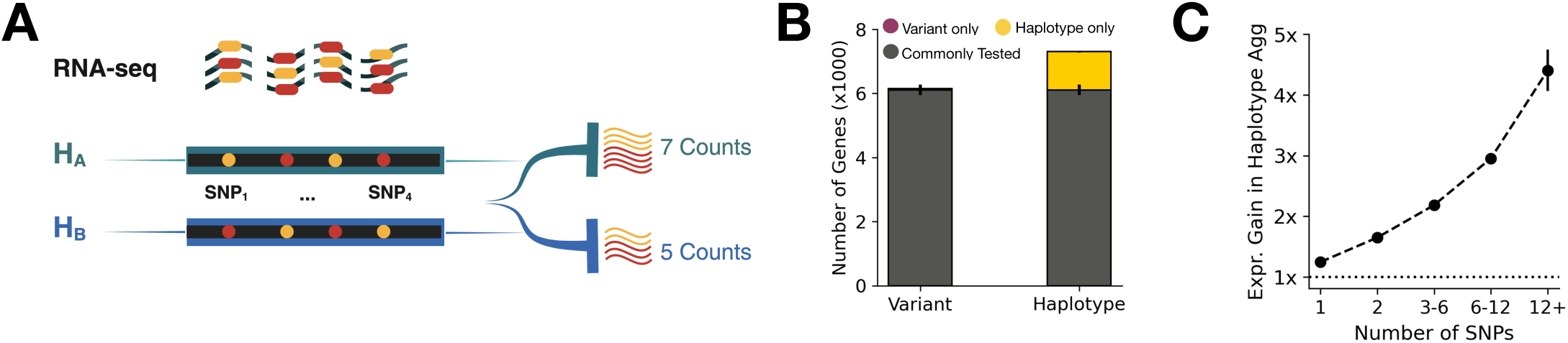
Allele-Specific Expression (ASE) Analysis and Haplotype Aggregation. **(A)** Schematic demonstrating haplotype aggregation of ASE reads in a RNA-seq library using SNPs in a phased haplotype block [21]. **(B)** Average number of protein-coding genes per sample with detectable ASE (total expression count > 10) exclusively at variant level, haplotype level, and those common to both across 49 GTEx tissues. Error bars represent the 95% confidence intervals (CI) of the average number of genes for each tissue. **(C)** Expression gain of ASE, calculated as the ratio of haplotype-level to variant-level total counts as a function of available ASE SNPs among commonly available protein coding genes. Error bars represent the 95% CI of the mean gain among 49 GTEx tissues.

### A Modified Generative Model for Haplotype-level ASE Data in Population

Variant-level ASE data in a population is described by a constrained mixture of binomial-logit-normal distribution as derived by Mohammadi et al. [8]. This model, ANEVA, is parametrized by the regulatory effect of the genetic variants in the population and their linkage disequilibrium to the SNP at which ASE is observed (aseSNP) and is used to estimate the genetic variance in gene expression (V^G^). Testing for dosage outliers from ASE data is implemented in the accompanying statistical outlier detection test, ANEVA-DOT, which uses V^G^ as a reference input informing of the scale of population variability expected in each gene. However, the haplotype-aggregation of ASE data obscures the linkage disequilibrium between the aseSNP and regulatory variants rendering the ANEVA model not appropriate for haplotype-level ASE data. Here we use a modified model catered to haplotype-level ASE data, ANEVA-h, where all regulatory effects are modeled within a single BLN distribution to estimate V^G^, expected variance in allelic expression due to genetic variation in regulation (See Supplementary Information). The V^G^ estimates from ANEVA-h are analytically consistent with those of ANEVA representing expected standard deviation of genetic regulatory variation in a gene in a population in log allelic fold change, and thus they are readily usable for downstream use in dosage outlier testing using ANEVA-DOT. Furthermore, ANEVA-h is computationally more efficient than the original ANEVA model, allowing for derivation of confidence intervals for V^G^ via parametric bootstrapping which is currently lacking (eq. 4, Supplementary Information). We provide ANEVA-h as a standalone R package or within a docker container to facilitate its application [22, 23].

### ANEVA-h Improves Expression Variation Analysis, Increases the Number of Tested Genes, and Reduces False Positives Across GTEx Data

We applied ANEVA-h on 15,201 samples from the GTEx V8 project release data and benchmarked it against those derived using the original ANEVA and reported in Ferraro et al. [8, 18]. We generated V^G^ estimates with confidence intervals, for 26,862 protein-coding and long-noncoding RNA (lncRNA) genes spanning 49 tissues from 838 individuals that had a minimum total ASE of 30 in at least six individuals per tissue (Table S1). Focusing on genes with estimates available in both, we compared V^G^ estimates from ANEVA-h to those generated using the original ANEVA, we observed a strong linear relationship between V^G^ of variant and haplotype level data, with a mean pearson correlation of 0.728 across 49 tissues (Table S1). To evaluate the biological relevance of these estimates, we next assessed the mean rank correlation of V^G^ values with measures of genic constraint and intolerance including pLI, RVIS, ncRVIS, and ncGERP across tissues. Compared to the original method, ANEVA-h V^G^ values showed consistently stronger rank-correlations with pLI, RVIS, and ncGERP compared to our previous variant-level estimates while correlations with ncRVIS remained unchanged (Fig S1).

Next, we assessed the number of genes for which V^G^ could be estimated using haplotype-versus variant-level data. The mean number of protein-coding genes with V^G^ data per tissue increased from 6,196 with ANEVA to 12,682 for ANEVA-h (2.04-fold gain; Fig. 2A-B, S2A). A total of 17,159 protein-coding genes had V^G^ estimates in at least one tissue, compared to 14,193 in variant-level analysis (Fig. 2C). Notably, the number of genes with available V^G^ estimates across all 49 GTEx tissues increased substantially from 328 in variant-level data to 7,521 in haplotype-level data (22.93-fold gain) (Fig. 2C). Subsequently, we analyzed the distribution of genes with V^G^ estimates, stratified by expression levels measured in transcripts per million (TPM). Across all expression levels, ANEVA-h consistently increased the number of protein-coding genes with available V^G^ estimates (Fig. 2C-D). However, the most pronounced gain was observed among genes with low expression levels (≤ 10 TPM), where the average number of genes with V^G^ estimates per tissue increased from 987 in the variant-level data to 4,768 (4.83-fold gain) in the haplotype-level data (Fig. 2C). Following this, we compared the number of genes with available V^G^s across a functionally diverse range of gene sets including autosomal dominant, dosage sensitive, haplo-insufficient, and loss-of-function (LoF) intolerant genes, that could lead to improved characterization of disease-relevant gene regulation and strengthen clinical diagnostic utility. (Table S1). We observed an over 2-fold average increase in genes with V^G^ estimates across all gene sets in haplotype-level data across all tissues (Fig. S2 D-E).

**Figure 2.**
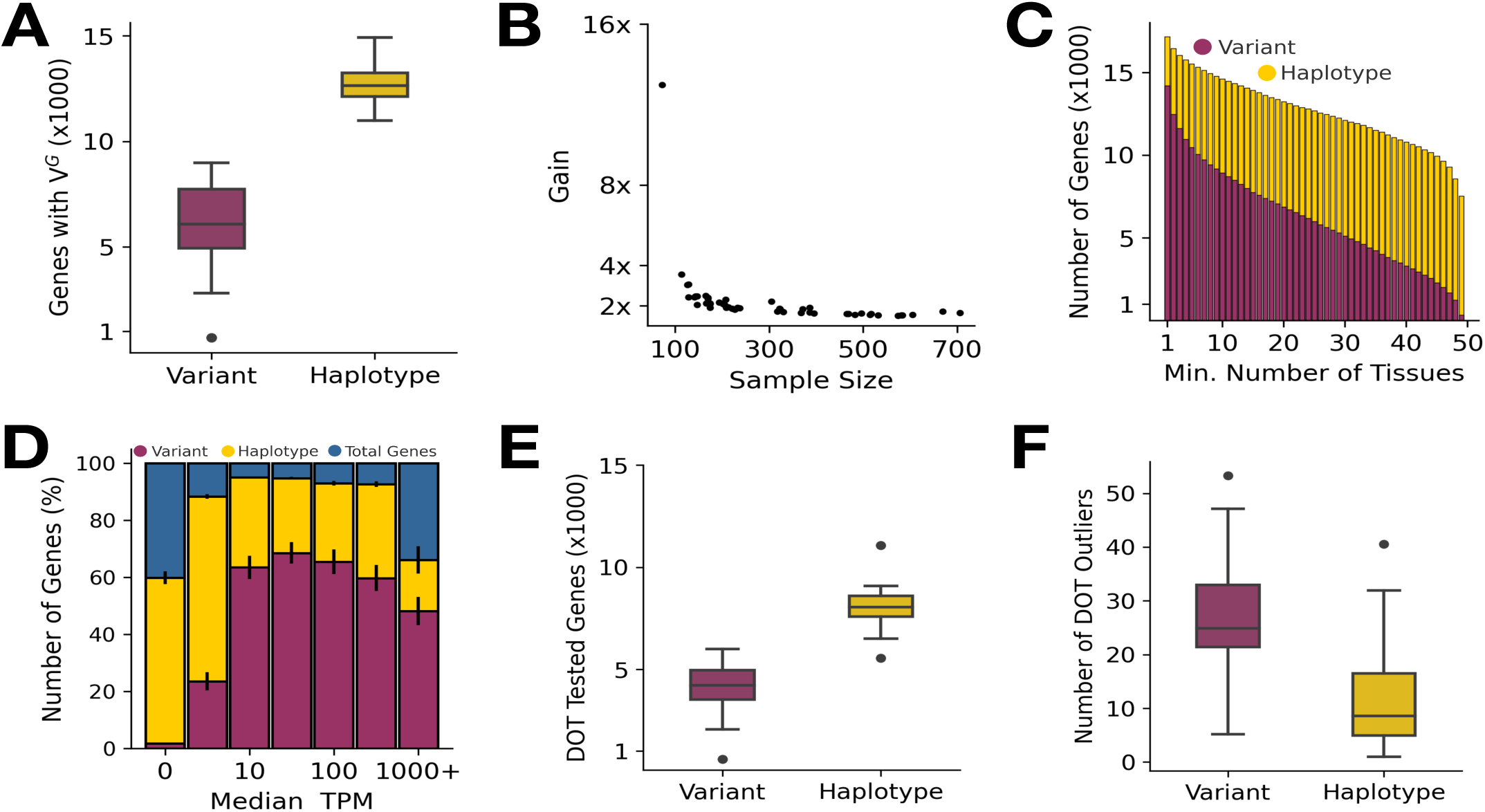
ANEVA-h provides accurate estimates of genetically regulated variation in gene expression in GTEx project data. **(A)** Distribution of genes with V^G^ estimates per GTEx tissue. **(B)** Increase in number of genes with V^G^s as a function of GTEx tissue sample size **(C)** total genes with V^G^ estimates across 1–49 GTEx tissues**. (D)** Average percentage of genes with V^G^ estimates by median TPM (normalized to bins). Error bars represent the 95% CI of the mean among 49 GTEx tissues. **(E-F)** Distribution of the average number of DOT Tested genes **(E)** and DOT outliers (q-value ≤ 0.05) per sample **(F)**.

Finally, we assessed the results of dosage outlier detection test. ANEVA-DOT was applied to all available samples across 49 GTEx tissues once using V^G^s derived from haplotype-level ANEVA-h and once using those from the original variant-level ANEVA [8]. On average, we observed that the number of tested protein coding genes per-sample rose from 4182 to 8064 (1.92-fold increase) across tissues (Fig. 2E, S3A).

Subsequently, we calculated the average number of significant outlier genes per sample having a q-value ≤ 5% (FDR adjusted; Benjamini-Hochberg). On average per-tissue, we observed that the mean number of outlier genes per sample reduced from 27.49 in variant level data to 11.23 (2.48-fold decrease) (Fig. 2F, S3B).

### ANEVA-h Outliers Have Higher Enrichment for Large-effect Rare Variants in GTEx

To evaluate whether ASE outliers are enriched for functionally impactful rare variants, we quantified the enrichment of SNVs with minor allele frequency (MAF) <0.1% in both GTEx and gnomAD located within 10kb upstream of the transcription start site (TSS) or within the gene body [18, 24]. For each gene identified as an ASE outlier in at least one individual, we aggregated all nearby rare variants observed across both outlier and non-outlier individual-gene pairs and annotated them using VEP. We then computed a relative risk statistic, defined as the odds ratio of an outlier individual carrying a rare variant of a given consequence class compared to a non-outlier. This framework allowed us to quantify the enrichment of rare, putatively disruptive variants near ASE outliers across models and VEP variant annotation classes.

Using this approach, we first benchmarked rare variant enrichment performance across multiple ASE outlier detection methods—namely ANEVA-h, ANEVA, DROP-MAE, beta-binomial, and binomial models—by comparing outliers identified across 581 GTEx adipose subcutaneous tissue samples. Among these methods, ANEVA-h consistently showed the strongest enrichment across several high-impact variant classes—such as splice donor, splice acceptor, and stop-gained variants—followed by ANEVA (Fig S3A). These results indicate that haplotype-level ASE aggregation more effectively prioritizes functionally consequent rare regulatory variant than extant approaches.

We then extended this comparison between ANEVA-h and ANEVA across all GTEx tissues. Using the same relative risk metric across genes tested by each method, we observed significantly stronger rare variant enrichment in haplotype-level data, stratified by both outlier significance and VEP consequence (Fig. 3B-C, Fig. S4A-AW, S5A-AW, [8, 12]). To ensure consistency with our prior benchmarks, we applied our previously described false-positive-prone gene filter based on a Clopper-Pearson confidence interval method [8, 12]. Notably, false-positive prone genes, previously shown to have inflated false-positive rates due to reliance on a single ASE SNP were nearly absent from the haplotype-level data across all tissues. Specifically, it decreased from an average of 165.97 false positive prone genes per tissue in variant-level data, to just 0.71 false-positive prone genes per tissue in haplotype aggregated data (Fig. 3D, Table S2, [8, 25]). These observations underscore the robustness of haplotype aggregation, particularly in genes with multiple informative ASE SNPs.

**Figure 3.**
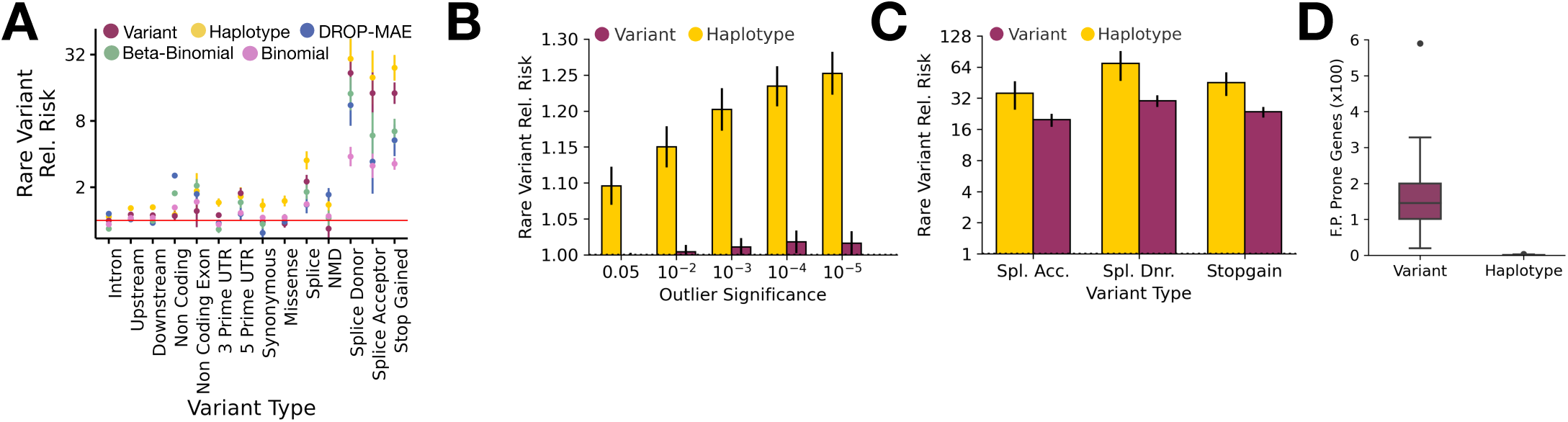
Improvements in Rare Variant Enrichment from Haplotype Aggregation in GTEx. **(A)** Relative risk estimates for rare variants (MAF < 0.1%) stratified by Variant Effect Predictor (VEP) consequence, comparing five models: Variant-level ANEVA, Haplotype-level ANEVA-h, DROP-MAE, Beta-Binomial, and Binomial models, in GTEx v8 adipose subcutaneous tissue **(B-C)** Rare variant relative risk stratified by statistical significance (FDR-corrected) **(B)** and VEP outlier classification **(C)** in variant-level and haplotype-level data. **(D)** Number of false-positive (F.P.) prone genes excluded per tissue in variant-level and haplotype-level ASE analyses across 49 GTEx tissues.

**Figure 4.**
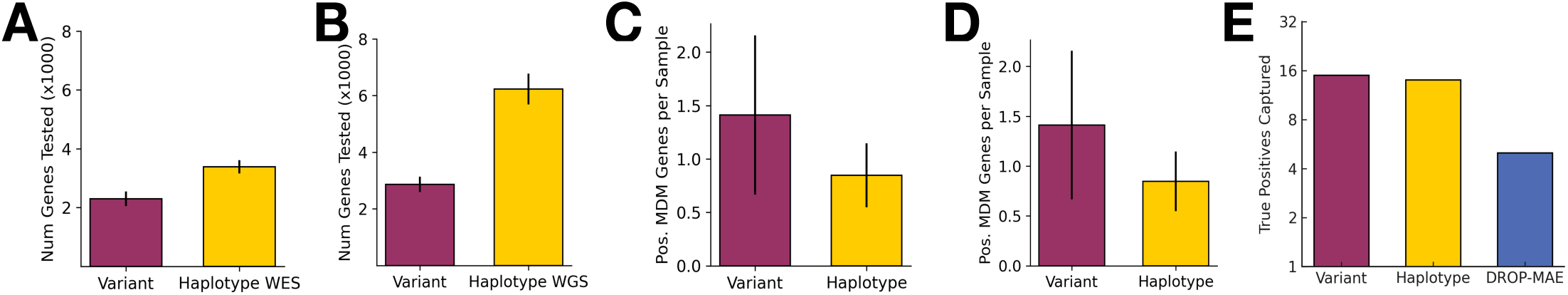
ANEVA-h Improves Diagnostic Utility in Rare Inherited Muscle Disease Cases (A-E) Analysis in muscular disease cohorts: **(A)** Number of genes tested per sample with whole-exome sequencing (WES); **(B)** Number of genes tested per sample with whole-genome sequencing (WGS); **(C)** Positive MDM genes identified per sample; **(D)** ANEVA-DOT test positivity rates. **(E)** Number of true-positive cases captured across variant, haplotype and DROP-MAE pipelines

Finally, we repeated the rare variant enrichment analysis after excluding false-positive prone genes from variant-level data. Even after this correction, haplotype-level models continued to show modest but consistent gains in rare variant enrichment compared to variant-level data, across both significance thresholds and functional VEP annotation classes (Fig. S6, S7A-AW, S8A-AW). These results demonstrate that haplotype aggregation improves the quantification of cis-regulatory variation, ultimately enhancing the ability to detect large-effect rare variants.

### ANEVA-h Enhances Transcriptome Interpretation in Rare Disease Cohorts

To evaluate the clinical utility of haplotype-based modeling of gene expression variance (V^G^), we applied ANEVA-h to two distinct rare disease cohorts including inherited muscular dystrophy and pediatric congenital heart disease (CHD). In the muscular dystrophy cohort (n = 46), comprising 18 genetically diagnosed cases and 28 unresolved cases, we generated haplotype-aggregated allele-specific expression (ASE) profiles using the PAC pipeline. Outlier detection was performed using ANEVA-DOT with V^G^ estimates derived from 706 skeletal muscle (MSCLSK) samples in GTEx v8. Compared to variant-level aggregation, haplotype-level aggregation increased the number of genes tested per sample by 1.47-fold for whole-exome sequencing (WES; from 2,285 to 3,372 genes) and 2.17-fold for whole-genome sequencing (WGS; from 2,866 to 6,231 genes) (Fig S4A-B). However, this increase in gene coverage was accompanied by a modest reduction in ANEVA-DOT test positivity rates—from 1.23% to 0.78% for WES and from 0.41% to 0.33% for WGS.

To benchmark the performance of ANEVA-h, we compared it against variant-level ANEVA and the recently published DROP-MAE pipeline across the 18 positive-control cases with known causal genes exhibiting ASE [7, 8]. Variant-level ANEVA identified 15 (83.3%), ANEVA-h identified 14 (77.7%), and DROP-MAE identified 5 (27.7%) (Fig S4E). Thirteen cases were shared between the ANEVA and ANEVA-h, and all five true positive cases identified by DROP-MAE calls were recovered by ANEVA and/or ANEVA-h. While variant-level ANEVA recovered the largest number of true positive cases, ANEVA-h improved the statistical significance of the causal gene in 9 of the 13 shared cases, indicating improved signal refinement (Table S3). Notably, ANEVA-h uniquely identified one diagnostic case (Sample D1) involving a large intergenic deletion in *LARGE1* that was missed by variant-level analysis (Fig S10).

Next, we assessed the performance of ANEVA and ANEVA-h on 28 undiagnosed cases (Table S3). The average number of significant MDM-panel genes per sample decreased from 1.47 (variant-level) to 0.29 (haplotype-level), with largely concordant results between methods. Importantly, in one case (Sample N19) we identified a novel candidate gene for further investigation (Table S3).

Next, we applied ANEVA-h to 257 samples from the Pediatric Congenital Heart Disease Consortium (PCGC), using GTEx heart tissues to match each sample to its closest transcriptomic reference: 64 samples to aorta (ARTAORT), 100 to atrial appendage (HRTAA), and 94 to left ventricle (HRTLV) [11, 17]. This yielded an average of 3,731, 3,522, and 3,365 genes tested per sample, respectively, with corresponding ANEVA-DOT test positivity rates of 0.51%, 0.64%, and 0.73% (Fig S11A, C). One sample was excluded from further analysis due to an abnormally high number of outliers. To validate our approach, we compared ANEVA-DOT findings to previously reported ASE events in 45 PCGC cases. Among the 38 overlapping samples, ANEVA-DOT rediscovered 21 (55.2%) of the previously observed events, confirming high concordance. To assess potential pathogenicity, we used the RENOVO framework to flag rare, deleterious variants in genes with significant ASE. Among all PCGC cases, 216 (84%) had no such variants, while the remaining 41 samples harbored an average of 1.317 candidate genes per sample (Fig S11B). Focusing on a curated panel of 396 CHD-associated genes, an average of 0.71 per sample showed both significant ASE and a likely pathogenic RENOVO variant. Two of these cases showed strong convergence between ASE outliers and known CHD gene variants and were prioritized for further clinical review.

Together, these findings demonstrate that ANEVA-h enhances transcriptome-based interpretation across diverse rare disease cohorts. By improving gene coverage and refining ASE signal detection, ANEVA-h facilitates both recovery of known pathogenic genes and identification of novel candidate gene–variant pairs for diagnosis and follow-up in unsolved cases.

### ANEVA-h Identifies Ancestry-specific Variability in Population ASE data

To evaluate how population structure influences estimates of regulatory variation, we generated ANEVA-h V^G^s and their 95% confidence intervals for each population group using allele-specific expression (ASE) data from a subset of MAGE study comprising of 701 individuals spanning 25 diverse populations (Table S5, [19, 26, 27]). Next, we computed a distance metric based on the overlap of confidence intervals to assess the similarity of V^G^ estimates across populations (See Methods). Hierarchical clustering of these distance metrics revealed distinct clusters that largely corresponded to major continental population groups—African (AFR), East Asian (EAS), European (EUR), and South Asian (SAS) (Fig. 5A).

**Figure 5.**
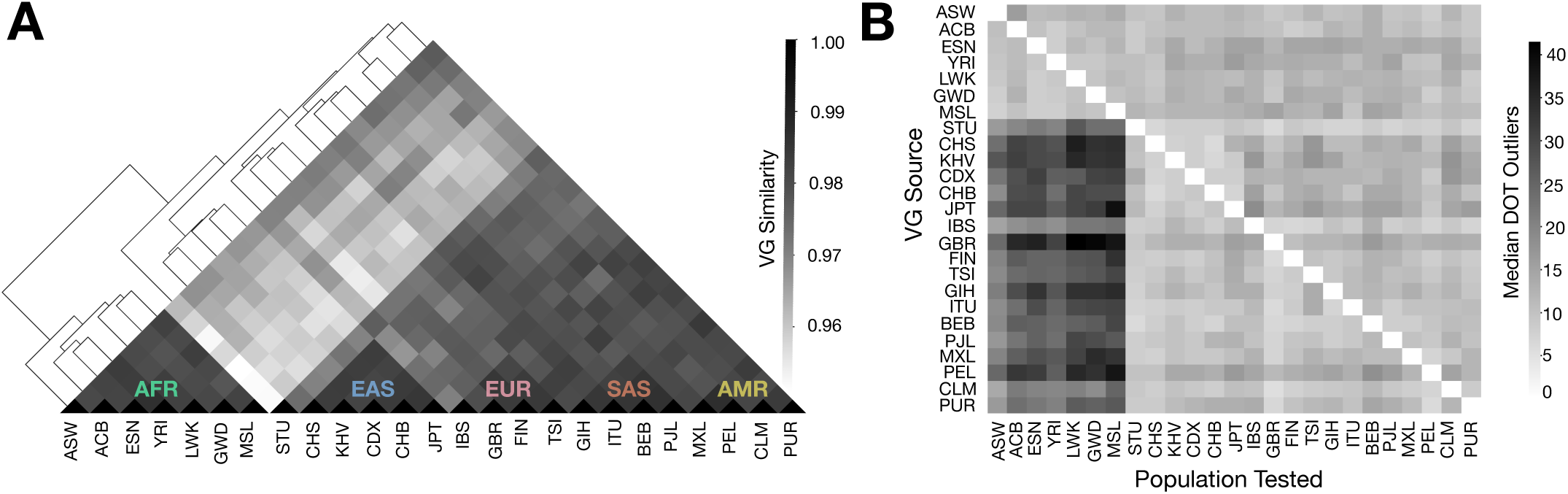
ANEVA-h Identifies Ancestry-Specific Variability in V^G^ Estimates. **(A)** Hierarchical clustering of ANEVA-h V^G^ confidence intervals across commonly tested genes from 25 population groups. Population codes correspond to standard abbreviations used within the 1000 genomes project. **(B)** Number of median ANEVA-DOT outliers across all tested ASE population-V^G^ combinations.

Next, we evaluated how mismatches in ancestry between the reference population and the tested individual affect ANEVA-DOT results. To do this, we systematically tested the ANEVA-DOT positivity rate using ASE data from every possible source population (Fig 5B). We found that when ASE data from African populations was tested against V^G^s derived from non-African groups, there was a 2.7-fold median increase in the number of ANEVA-DOT positive genes. In contrast, when non-African ASE data was tested against African-derived V^G^s, the effect was much more modest, corresponding to only a 1.2-fold median increase (Fig. 5B). This asymmetry could be suggestive of the higher genetic diversity and regulatory variation in African populations [28–30].

Next, we assessed the extent to which ANEVA-DOT positivity rates correlate with population genetic distance, as measured by the fixation index (FST), and observed a linear relationship between FST and the number of median DOT outliers, with the strongest pearson correlation seen (in African ancestry groups (Fig S12, [31]). These findings are similar to previous observation across the genomics field and underscore the challenges for application of ANEVA-DOT across population and the value of ancestry-matched resources for better quantification of regulatory variation [32].

## Discussion

Here, we introduced ANEVA-h, to enable use of haplotype-aggregated allele-specific expression to enhance the detection of large effect rare genetic regulatory variants from transcriptome sequencing data. By moving beyond single-SNP ASE measurements, ANEVA-h provides a more comprehensive approach to quantifying gene expression variance (V^G^), improving sensitivity while reducing false positives. This refinement enables more accurate prioritization of regulatory variants, strengthening the link between transcriptomic variation and genetic disease. One of ANEVA-h’s key advantages is its ability to detect ASE in a broader range of genes, particularly those with low expression, by capturing cumulative regulatory effects across multiple SNPs. This aggregation reduces false positives seen in ANEVA-DOT and improves ASE-based outlier detection. The enhanced enrichment for rare variants further underscores the biological relevance of detected ASE signals, reinforcing its value in identifying pathogenic regulatory effects. ANEVA-h also demonstrates improved performance in clinical datasets, refining gene prioritization and filtering out spurious associations in neuromuscular and cardiac disorders, thereby making ASE-based diagnostics more precise. Additionally, analyses from the MAGE cohort reveal ancestry-driven differences in V^G^ estimates, emphasizing the need for population-aware ASE analysis to improve accuracy in diverse cohorts. From a computational perspective, ANEVA-h simplifies statistical modeling, reducing computational complexity and the availability of a containerized implementation ensures scalability to larger datasets and enhances reproducibility across computing environments. From practical perspective, haplotype-aggregated ASE data needed for ANEVA-h, unlike variant-level ASE data, is easy to share and can be freely downloaded for GTEx and other reference datasets as it does not contain genetic data.

Despite these advancements, ANEVA-h has several limitations. Its reliance on phased genotypes introduces the risk of misclassifying or missing true regulatory signals, as demonstrated by two muscular dystrophy cases that were identified using our variant-level ANEVA approach but lost with ANEVA-h. Since the genotypes were population-phased, rare ASE-driving variants were dropped out during the phasing process, obscuring allele-specific signals. Trio phasing could mitigate these issues by providing direct inheritance patterns, but this remains challenging—particularly in cases involving de novo variants where parental genotypes offer no reference for phasing. Moreover, clinically, trios are often unavailable, further limiting the feasibility of this approach in routine diagnostics. Another limitation is ANEVA-h’s dependence on GTEx for V^G^ estimation, restricting its applicability to ancestrally diverse and disease-specific cohorts. Expanding reference datasets will be essential for improving the accuracy of ASE-based analyses across broader populations. Additionally, while ANEVA-h is highly effective in detecting dominant-acting regulatory effects, it is less informative for recessive variants, which require family-based analyses or complementary genomic approaches.

## Methods

### Variant-Level ASE and ANEVA-DOT on GTEx

For samples stratified by GTEx tissue, allele-specific expression (ASE) analysis was performed at the variant level using ANEVA-DOT [8, 18]. The ANEVADOT_test function in R was applied with default settings and a minimum coverage threshold of 10 reads. Reference variance of gene expression (V^G^) estimates were obtained from GTEx v8 and our previously published ANEVA-DOT reference datasets [8]. SNP-level ASE data was extracted from protected GTEx portal data and cross-referenced with sample metadata for tissue-specific stratification. For each gene, the SNP with the highest total expression was selected as the representative SNP for ANEVA-DOT analysis. False discovery rate (FDR) correction was applied per sample on protein-coding and long non-coding RNA (lncRNA) genes as defined in GENCODE v26 using the Benjamini-Hochberg method.

### Haplotype-Level ASE Analysis

#### Data Preparation for ANEVA-h on GTEx

Haplotype-aggregated ASE data was obtained from GTEx v8 haplotype-expression matrices. Sample metadata, including tissue type, was retrieved from the publicly available GTEx v8 sample annotation dataset [18]. Using this metadata, samples were stratified by tissue type, and input files of reference and alternative allele counts were generated.

#### V^G^ Generation Using ANEVA-h

For each GTEx tissue, V^G^ estimates and parametric confidence intervals were computed using the anevah_bootstrap.R script from the ase_outlier_tools container [23]. V^G^ estimation was performed using default model parameters with 1000 iterations for parametric bootstrap and model fitting executed using 16 CPU cores. V^G^ calculations were restricted to genes with a minimum total expression of 30 in at least six individuals per tissue. The resulting V^G^ estimates with confidence intervals for 49 GTEx tissues are available for download in our GitHub repository (https://github.com/PejLab/ASE_VG/), from Table S1, or on zenodo [26].

#### ANEVA-DOT from Haplotype-level ASE

ANEVA-DOT analysis was also performed on haplotype-level ASE data following the same methodology as for variant-level ASE. Briefly, the ANEVADOT_test function was applied with default settings and a coverage threshold of 10 reads. Reference V^G^ estimates were derived from haplotype-level data, and FDR correction was again applied to protein-coding and lncRNA genes as defined in GENCODE v26 using the Benjamini-Hochberg method.

#### Gene Expression Values

To assess the number of genes with V^G^ values for both variant and haplotype level ASE based on expression levels, transcript-per-million (TPM) values were extracted from the GTEx v8 dataset [18]. Samples were stratified by tissue type, and the median expression per gene was calculated across available samples per tissue. Protein-coding genes with a median TPM greater than 1.58 were retained for reporting statistics.

#### Gene Sets

Curated gene lists from our previous study were used to evaluate the number of genes with V^G^ estimates across various functional categories (Table S1). Additionally, we examined the number of genes genes predicted to exhibit haploinsufficiency and triplosensitivity, using predicted classifications from recent studies [24, 33]. Predictions were downloaded from these results and haploinsufficiency (pHaplo) and triplosensitivity (pTriplo) scores were categorized as strong (≥0.86 and ≥0.94, respectively), moderate (0.86–0.55 and 0.94–0.68), or weak (<0.55 and <0.68), in line with the classification thresholds used by DECIPHER [34].

#### Rare Variant Enrichment in GTEx

To examine the relationship between rare variants and ASE outliers, single nucleotide variants (SNVs) with a minor allele frequency (MAF) less than 1% were identified from GTEx genotype data and cross-referenced with gnomAD v2.0.2. Functional consequences of these variants were predicted using Variant Effect Predictor (VEP), and variants were assigned to genes if they were located within a ±10 kb window of gene boundaries [12]. Relative risk for rare variant enrichment was calculated using the ExOutBench framework in R, consistent with prior works [12]. Briefly, Relative risk was estimated by selecting genes with significant outlier scores in at least one individual, and odds ratios were computed to compare the likelihood of rare variants in ASE outlier versus non-outlier individuals. Enrichment scores were further refined by stratifying results across varying outlier significance thresholds and VEP annotation categories using the enrichment_by_significance and enrichment_by_annotations functions with default settings.

#### Clopper Pearson Method for Identifying False-positive Prone Genes in GTEx

To detect false-positive prone genes, the frequency of ANEVA-DOT outliers per gene was computed across GTEx tissues for both variant and haplotype level ASE. A binomial test was performed using the binom_test function from the scipy.stats module, and confidence intervals (ala clopper pearson exact method) were estimated using the proportion_ci utility [35]. Genes were classified as false-positive prone and added to exclusion lists if the lower bound of their confidence interval exceeded 1%. Final lists of such false-positive genes for both variant- and haplotype-aggregated ASE data are curated and available from our GitHub repository (http://github.com/PejLab/ASE_FP_Lists/), Table S2 or on zenodo [25].

#### Muscular Dystrophy Cohort

RNA and DNA sequencing data were obtained for 46 individuals from the Genetics of Inherited Muscle Disease cohort, available through the Database of Genotypes and Phenotypes (dbGaP; accession: phs000655.v3.p1). These individuals presented with a heterogeneous spectrum of neuromuscular disorders, including congenital muscular dystrophy, congenital myopathy, limb-girdle muscular dystrophy, Emery-Dreifuss muscular dystrophy, and arthrogryposis. To enable cross-study comparisons, case identifiers were mapped to existing hash IDs from prior analytical efforts [8, 9]. Only cases with publicly available sequencing data in dbGaP were included in this study, while cases previously unpublished and consequently not part of dbGaP were excluded.

#### Variant Level ASE Data and ANEVA-DOT

Allele-specific Expression Data (ASE) and ANEVA-DOT results were downloaded from our previously published effort for the corresponding cases and used for subsequent comparisons with haplotype-level ASE data [8]. Subsequent downstream analysis involved reporting outlying protein-coding genes, with a particular emphasis on a curated set of 193 muscular dystrophy-relevant genes used from our previous efforts (Table S3, [8]).

#### Haplotype Level Data Sequencing Data Processing

Both RNA-sequencing (RNA-seq) and proband DNA-sequencing data were retrieved and subjected to standardized and quality control measures. The specific analytical pipelines used for RNA-seq and DNA-seq processing are described in subsequent sections.

#### RNA-seq Data

Paired-end RNA-seq data was downloaded from dbGAP and quality control for RNA-seq reads was performed using FASTQC [36].

#### DNA-seq Data

Genotypes, derived from either whole-exome sequencing (WES) or whole-genome sequencing (WGS), were processed using a standardized pipeline. Briefly, genotype coordinates were converted from the human reference genome hg19 (GRCh37) to hg38 (GRCh38) using the CrossMap utility in Python 3.8.3 [37]. Following liftover, genotypes were separated by chromosome using bcftools, generating individual chromosome-specific VCF files [38]. Each chromosome-specific VCF was population phased using the EAGLE, using reference maps from the 30× Illumina NovaSeq sequencing dataset of the 1000 Genomes Project of high-coverage phased genotypes from 3,202 individuals [39]. The phased genotype files were subsequently merged using Picard MergeVcfs, followed by sorting and indexing with bcftools and tabix to generate the final phased VCF file for each sample [40–42].

#### Allele-specific Expression (ASE) Data Generation

Allele-specific expression (ASE) data were produced using the Personalized Allele Caller (PAC) pipeline [16]. The Nextflow workflow was executed using default parameters with GRCh38 as the reference genome and Gencode v26 as the annotation reference [43]. Briefly, the pipeline involves multiple stages to quantify ASE at the gene level from both maternal and paternal genomes. STAR (v2.7.4a) was used in two-pass mode to align paired-end RNA-seq reads to the reference genome [44]. Local phasing of genotypes was performed using pHASER (v1.0.0), after which phased variants were used to construct personalized maternal and paternal diploid genomes via VCF2Diploid (v0.2.6a) [15, 45]. Reads were separately mapped to these genomes to derive allele-specific read counts. To account for multimapping reads, RSEM was employed for probabilistic assignment based on transcript abundance estimates [46]. Phaser Gene AE was then used to quantify haplotypic read counts for each allele independently, and final allele-specific expression counts were aggregated at the gene level from both genomes [15]. The pipeline was executed on a high-performance computing (HPC) node using 16 CPU cores and 185GB of RAM via a SLURM job scheduler.

#### ANEVA-DOT for MDM Cases

ASE data for each sample was produced using the workflow as outlined above. Haplotype-level V^G^ estimates were generated by running ANEVA-h on GTEx v8 skeletal muscle samples, following the procedures detailed in previous sections. Subsequently, the ANEVADOT_test function from the ANEVA-DOT R package was applied using default parameters, with a minimum coverage threshold of 10 reads to compute gene-level p-values for each sample. In accordance with prior approaches, raw p-values were adjusted for false discovery rate (FDR) using the Benjamini-Hochberg method, restricting the analysis to protein-coding and long non-coding RNA (lncRNA) genes as defined in Gencode v26. Finally, consistent with variant-level data, downstream analysis focused on identifying outlying protein-coding genes, with particular emphasis on a curated set of 193 muscular dystrophy-relevant genes from our previous efforts (Table S3, [8]).

#### DROP-MAE on Positive Control MDM Cases

RNA sequencing (RNA-seq) reads were aligned to the human reference genome (GRCh38) using STAR v2.7.9a in two-pass mode, followed by sorting and indexing with Samtools v1.15. Phased genotypes were generated as previously described. The DROP monoallelic expression (mae) module was executed using snakemake with gencode v26 annotations and GRCh38 specified in the configuration file [7, 47]. To ensure consistency with variant-level analyses, DROP outputs were processed as follows: ASE counts were aggregated at the gene level by selecting the SNP with the highest expression as the representative SNP for each gene, in accordance with the variant-level methodology described earlier; raw p-values from gene-level ASE tests were filtered to include only protein-coding and long non-coding RNA (lncRNA) genes; and multiple testing correction was performed using the Benjamini-Hochberg false discovery rate (FDR) adjustment. The resulting FDR-adjusted p-values were used to benchmark results against positive control cases.

#### Congenital Heart Dystrophy Cohort Cohort Description

The Pediatric Cardiac Genomics Consortium (PCGC) launched the CHD GENES study (NCT01196182) to collect genotype and phenotype data from a large congenital heart disease (CHD) cohort [17]. Probands were recruited from five primary sites (CHOP, Columbia, Harvard/Boston Children’s, Mount Sinai, and Yale) and four satellite sites (CHLA, Cohen Children’s, UCL, and University of Rochester) in the US and UK, beginning in November 2010. Recruitment protocols varied by site, but all CHD diagnoses were confirmed via imaging and operative reports, coded using the International Paediatric and Congenital Cardiac Codes, and manually reviewed by investigators. Parental and clinical data, including genetic tests, physical exams, and extracardiac anomalies, were collected through interviews and medical records. Our study uses a subset of CHD GENES cases with available heart tissue. From 397 surgery-discarded tissue samples from 350 patients, we retained 257 samples from 220 patients after excluding those lacking matching RNA-seq and genotype data or with >10% missing genotypes. Tissues were categorized according to GTEx conventions as Atrial Appendage (HRTAA, n=100), Aorta (ARTAORT, n=64), and Left Ventricle (HRTLV, n=94) (Table S4). Most patients were infants, with a median age at consent of 6 months (range: 0–21 years).

#### RNA-seq data generation

Total RNA was extracted from RNAlater-preserved frozen cardiac tissue using TRIzol Reagent (Life Technologies) according to the manufacturer’s instructions. Samples with RNA integrity number (RIN) > 5 were selected for sequencing. Polyadenylated RNA was enriched via two rounds of oligo(dT) selection, converted to cDNA, and used to generate RNA-seq libraries following standard protocols. Libraries were sequenced on Illumina HiSeq 2000 or HiSeq 2500 platforms to produce 50 bp paired-end reads, targeting >20 million reads per sample (median: 57 million). Mitochondrial reads were excluded using Samtools. Initial alignments to the GRCh37/hg19 reference genome were performed by the PCGC consortium using TopHat v1.4 [48]. To enable downstream analyses which requires GRCh38/hg38, we re-aligned all RNA-seq data to the hg38 reference genome. BAM files were reverted to unaligned BAMs using Picard RevertSam (v2.21.3), converted to FASTQ format using the bamtofastq utility from bedtools v2.25.0, and realigned using STAR v2.7.3 in two-pass mode with the --waspOutputMode SAMtag option enabled to correct for allele-specific mapping bias using the WASP framework [49, 50].

#### WES and WGS data generation

Whole-exome sequencing (WES) was performed at the Yale Center for Genome Analysis, as previously described [51]. Briefly, genomic DNA (gDNA) extracted from venous blood was captured using the NimbleGen v2.0 exome capture kit (Roche) and sequenced on the Illumina HiSeq 2000 platform with 75 bp paired-end reads to a mean depth of 107×. Reads were aligned to the GRCh37/hg19 reference genome using Novoalign (Novocraft) and processed following GATK Best Practices workflows [52]. Variant calling for single nucleotide variants (SNVs) and indels was performed using GATK HaplotypeCaller [53]. Whole-genome sequencing (WGS) was carried out at the Baylor College of Medicine Genomic and RNA Profiling Core, the New York Genome Center (NYGC), and the Broad Institute. gDNA from venous blood or saliva was used to generate PCR-free libraries, which were sequenced on the Illumina HiSeq X Ten platform with 150 bp paired-end reads, achieving a median coverage >30× per sample. Reads were aligned to either GRCh37/hg19 or GRCh38/hg38 using BWA-MEM, and processed following GATK Best Practices, including base quality score recalibration, indel realignment, and duplicate marking [42, 52]. VCF files were reviewed by clinical genetics experts at New York-Presbyterian Hospital/Columbia University Medical Center. All sequencing and variant review procedures were conducted by the PCGC consortium.

#### WES and WGS preprocessing

VCF files were lifted over from the Hg19 to Hg38 build of the human genome using Crossmap (v.0.4.2) [37]. 70 patients were sequenced twice, once as WES and a second time as WGS. For these patients, both files were concatenated into a unique file, dropping duplicate records. Then, the number of missing genotypes calls per patient was assessed and patient’s sex was empirically confirmed as part of our quality control check using Plink v2.00, an open-source whole genome association analysis toolset [54]. Patients having more than 10% missing genotype calls were removed from downstream analysis. VCF files were then phased for each patient using the 1000 genomes dataset as a reference panel +/- parents information when it was available using SHAPEIT4 [55].

#### ASE data generation and ANEVA-DOT Outlier Test

We assessed potential identity mismatches between VCF files and RNA-seq data using the MBV utility in QTLtools v1.2, correcting inconsistencies where appropriate [56]. Allele-specific expression (ASE) counts were generated from uniquely mapping reads using phASER using RNA-seq BAM files, filtered to retain only reads passing WASP quality criteria [15, 49]. The resulting ASE data was annotated using GENCODE v26. We then applied ANEVA-DOT using V^G^ estimates from ANEVA-h, selecting the appropriate heart tissue (ARTAORT, HRTAA, HRTLV) for each sample, with default settings and a minimum coverage threshold of 10 reads. To identify ASE outliers, FDR correction was performed for each sample on a subset of protein-coding and lncRNA genes, as defined in GENCODE v26.

#### Variant Prioritization Using Renovo

Among ASE outlying genes identified through ANEVA-DOT, we prioritized putative deleterious variants exhibiting demonstrable ASE using ReNOVo [57]. Variants with ASE, as determined from phASER outputs, were first subset using bcftools [38]. ReNOVo was then applied using default settings to ascertain putative deleterious variants [57]. Functional annotation for ReNOVo was performed using ANNOVAR with annotation for the hg38 genome build [58]. From the predictions, variants labeled with a pathogenic flag (low precision LP, intermediate precision IP, and high precision HP) within the RENOVO_Class field were subsequently prioritized for further analysis.

#### MAGE Cohort

The MAGE cohort consists of 731 individuals selected from the 1000 Genomes Project, representing 26 diverse population groups, with approximately 30 individuals per group [19]. This dataset includes genomic and transcriptomic data derived from Lymphoblastoid Cell Line (LCL) samples. We chose to analyze 701 individuals from 25 population groups excluding Utah residents (CEPH) with Northern and Western European ancestry (CEU) population group.

#### RNA-seq Data

Paired-end RNA-seq data for 731 samples were downloaded from the Sequence Read Archive (SRA) using sra-toolkit (v3.0.10) (Accession: PRJNA851328) [19]. Quality control of the FASTQ files was performed using FastQC to assess read quality and sequencing artifacts [36]. For both datasets, alignment of RNA-seq reads was performed using STAR (v2.7.9a) using two-pass mode (--twopassMode Basic) against the GRCh38 reference genome (GCA_000001405.15), excluding alternate loci and decoys [44]. BAM files were generated, coordinate-sorted, and indexed using samtools (v1.10) [38].

#### Genotypes

Phased genotypes were downloaded from the 30x Illumina NovaSeq study by the 1000 Genomes Consortium High-coverage phased genotypes (https://ftp.1000genomes.ebi.ac.uk/vol1/ftp/data_collections/1000G_2504_high_coverage/working/20220422_3202_phased_SNV_INDEL_SV/) [39]. Genotyping information for both Contamination and MAGE datasets were processed through a standardized pipeline: Chromosome-level VCFs were first split by sample using bcftools view (v1.10.2), merged with Picard MergeVcfs (v2.27.1), and then sorted and indexed using bcftools and tabix [38, 40, 41].

#### Allele-Specific Expression Data

ASE data was produced using phASER (v1.0.0) with best-practice parameters, and genomic region blacklisting for GRCh38. Haplotype counts were extracted from haplotypic_counts.txt files and summarized using the phaser_gene_ae tool (Python v2.7.11) [15]. Aggregated ASE data were mapped to protein-coding genes using gencode v26 annotations. ASE Matrices derived from the MAGE cohort are provided at https://github.com/PejLab/ASE_Data or archived on zenodo [27, 59].

#### V^G^ Generation Using ANEVA-h

V^G^ estimates and their corresponding parametric confidence intervals were computed separately for each of the 25 population groups using the anevah_bootstrap.R script from the container [23]. V^G^ estimation was performed using default model parameters, with 1000 iterations for parametric bootstrapping and model fitting executed using 16 CPU cores to optimize computational efficiency. To ensure statistical reliability, V^G^ calculations were restricted to genes with a minimum total expression of 30 in at least six individuals per tissue. The final V^G^ estimates, along with confidence intervals for all 25 ancestry groups, are available from our GitHub repository (https://github.com/PejLab/ASE_VG/), Table S5 or on zenodo [26].

#### Distance Metric for Identifying Ancestry-specific Variability in V^G^s

To assess similarities in V^G^ estimates across populations, we computed a distance metric based on confidence interval overlap and applied hierarchical clustering to identify ancestry-driven patterns. We first identified common genes present in all populations by taking the intersection of gene lists from each population’s V^G^ output file, retaining only these genes for downstream analyses. For each common gene, we evaluated confidence interval overlap across all pairwise population comparisons. V^G^ data from each population was extracted, and a binary overlap matrix was constructed, where each population pair received a value of 1 if their confidence intervals overlapped and 0 otherwise. To derive a distance metric reflecting V^G^ similarity, pairwise overlap scores were aggregated across all common genes and normalized by the total number of common genes to ensure consistency across populations. Under this normalization, a value of 1 indicates complete overlap of V^G^ confidence intervals across all common genes, whereas a value of 0 indicates no overlap. Finally, this distance metric was computed for all 25 population pairs and visualized using hierarchical clustering with the seaborn clustermap function under default settings [60].

#### ANEVA-DOT Across Population Groups in MAGE

To assess the impact of reference V^G^ estimates on ANEVA-DOT outlier rates across different population groups, ASE data from each population was systematically tested against V^G^ estimates from all other populations using the ANEVADOT_test function in the ANEVADOT R library with default settings and a coverage cutoff of 10 reads. FDR-adjusted p-values were computed, and samples with an adjusted p-value < 5% were identified as ANEVA-DOT outliers. The median number of DOT outliers per sample was then calculated for each ASE–V^G^ combination. To evaluate the impact of using reference V^G^ estimates from genetically divergent population groups on ASE outlier detection, mean fixation index (FST) values were computed using vcftools for genotypes from chromosome 21 on samples in each population pair to quantify genetic divergence [61]. The median number of DOT outliers using each reference V^G^s from each population group was then compared, with the mean FST values used as a measure of genetic distance between these groups to assess how divergence influences ASE outlier detection rates.

#### Contributions

K.G. led the scientific investigation, performed all analyses, interpreted results, and co-wrote the manuscript. M.B. co-led the study and contributed to data generation and analysis for the congenital heart disease cohort. S.S. provided clinical interpretation for the muscular disease analyses. M.M., E.S., B.K., and P.H. contributed to statistical model development and software implementation. A.T. provided analytical input and support. C.B., D.R.A, and T.L. provided conceptual input and analytical guidance. P.M. conceived and designed the study, supervised the project, secured funding, and co-wrote the manuscript. All authors reviewed and approved the final manuscript.

## Data and Code Availability

The allele-specific expression (ASE) data analyzed for this study for GTEx are available to authorized users through dbGaP under accession no. phs000424.v8 and on the GTEx portal (https://gtexportal.org/). The statistical model and code underlying ANEVA-h is available for download at https://github.com/PejLab/ANEVAh_repo as an R package or within a container https://hub.docker.com/repository/docker/krganapathy/ase_outlier_tools/. Reference V^G^ estimates and lists of false-positive prone genes from GTEx V8 across all 49 Tissues are available from Table S1 or available on GitHub https://github.com/PejLab/ASE_VG/and https://github.com/PejLab/ASE_FP_Lists.

Gene-level ASE Data from the Muscular Dystrophy and PCGC Heart Disease cases are available from Tables S3 and Table S4. All datasets and code, excluding those derived from clinical case studies, are publicly available via zenodo [22, 23, 25, 26].

## Data Availability

The allele-specific expression (ASE) data analyzed for this study for GTEx are available to authorized users through dbGaP under accession no. phs000424.v8 and on the GTEx portal (https://gtexportal.org/home/). The statistical model and code underlying ANEVA-h is available for download at https://github.com/PejLab/ANEVAh_repo as an R package or within a container https://hub.docker.com/repository/docker/krganapathy/ase_outlier_tools/. Reference VG estimates and lists of false-positive prone genes from GTEx V8 across all 49 Tissues are available from Table S1 or available on GitHub https://github.com/PejLab/ASE_VG/ and https://github.com/PejLab/ASE_FP_Lists. Gene-level ASE Data from the Muscular Dystrophy and PCGC Heart Disease cases are available from Tables S3 and Table S4. All datasets and code, excluding those derived from clinical case studies, are publicly available via zenodo.

## Acknowledgements

We gratefully acknowledge JC Ducom, Lisa Dong, and the Scripps Research High Performance Computing team for their exceptional computational support, as well as Douglas Evans, Salvatore Loguercio, Nathan Wineinger, Chunlei Wu, Andrew Su, and Xin Jin for their insightful discussions and critical feedback throughout this work. We recognize the Pediatric Cardiac Genomics Consortium (PCGC) for generating and providing access to essential datasets under the National Heart, Lung, and Blood Institute’s Bench to Bassinet Program (https://benchtobassinet.com), supported through grants UM1HL128711, UM1HL098162, UM1HL098147, UM1HL098123, UM1HL128761, and U01HL131003; this manuscript was prepared in collaboration with PCGC investigators and has been reviewed and approved by the consortium, whose members are listed at https://benchtobassinet.com/Centers/PCGCCenters.aspx. We also extend our gratitude to the GTEx donors and their families for their indispensable contributions to biomedical research, and acknowledge the Laboratory, Data Analysis, and Coordinating Center (LDACC) and the Analysis Working Group (AWG) for their efforts in generating and curating this foundational resource.

## References

1. Cleary, S. and C. Seoighe, Perspectives on allele-specific expression. Annual Review of Biomedical Data Science, 2021. 4: p. 101–122.

2. Lappalainen, T., et al., Transcriptome and genome sequencing uncovers functional variation in humans. Nature, 2013. 501(7468): p. 506–511.

3. Yépez, V.A., et al., Clinical implementation of RNA sequencing for Mendelian disease diagnostics. Genome medicine, 2022. 14(1): p. 38.

4. Byron, S.A., et al., Translating RNA sequencing into clinical diagnostics: opportunities and challenges. Nature Reviews Genetics, 2016. 17(5): p. 257–271.

5. Mohammadi, P., et al., Quantifying the regulatory effect size of cis-acting genetic variation using allelic fold change. Genome research, 2017. 27(11): p. 1872–1884.

6. Gasperskaja, E. and V. Kučinskas, The most common technologies and tools for functional genome analysis. Acta Medica Lituanica, 2017. 24(1): p. 1.

7. Yépez, V.A., et al., Detection of aberrant gene expression events in RNA sequencing data. Nature Protocols, 2021. 16(2): p. 1276–1296.

8. Mohammadi, P., et al., Genetic regulatory variation in populations informs transcriptome analysis in rare disease. Science, 2019. 366(6463): p. 351–356.

9. Cummings, B.B., et al., Improving genetic diagnosis in Mendelian disease with transcriptome sequencing. Science translational medicine, 2017. 9(386): p. eaal5209.

10. Kremer, L.S., et al., Genetic diagnosis of Mendelian disorders via RNA sequencing. Nature communications, 2017. 8(1): p. 15824.

11. McKean, D.M., et al., Loss of RNA expression and allele-specific expression associated with congenital heart disease. Nature communications, 2016. 7(1): p. 12824.

12. Ferraro, N.M., et al., Transcriptomic signatures across human tissues identify functional rare genetic variation. Science, 2020. 369(6509): p. eaaz5900.

13. Pastinen, T., Genome-wide allele-specific analysis: insights into regulatory variation. Nature Reviews Genetics, 2010. 11(8): p. 533–538.

14. Buil, A., et al., Gene-gene and gene-environment interactions detected by transcriptome sequence analysis in twins. Nature genetics, 2015. 47(1): p. 88–91.

15. Castel, S.E., et al., Rare variant phasing and haplotypic expression from RNA sequencing with phASER. Nature communications, 2016. 7(1): p. 1–6.

16. Saukkonen, A., H. Kilpinen, and A. Hodgkinson, Highly accurate quantification of allelic gene expression for population and disease genetics. Genome Research, 2022. 32(8): p. 1565–1572.

17. Hoang, T.T., et al., The congenital heart disease genetic network study: cohort description. PloS one, 2018. 13(1): p. e0191319.

18. Consortium, G., The GTEx Consortium atlas of genetic regulatory effects across human tissues. Science, 2020. 369(6509): p. 1318–1330.

19. Taylor, D.J., et al., Sources of gene expression variation in a globally diverse human cohort. Nature, 2024: p. 1–9.

20. Castel, S.E., et al., Tools and best practices for data processing in allelic expression analysis. Genome biology, 2015. 16(1): p. 1–12.

21. Ganapathy, K. Allele-Specific Expression (ASE) Analysis and Haplotype Aggregation. 2025; Available from: https://BioRender.com/45on04l.

22. Song, E., P. Hoffman, and P. Mohammadi ANEVA-h R Package. 2025. DOI: 10.5281/zenodo.15226575.

23. Ganapathy, K.R. and P. Mohammadi Singularity Container of Tools for Allele-Specific Expression Analysis. 2025. DOI: 10.5281/zenodo.15226244.

24. Karczewski, K.J., et al., The mutational constraint spectrum quantified from variation in 141,456 humans. Nature, 2020. 581(7809): p. 434–443.

25. Ganapathy, K.R. and P. Mohammadi False Positive Prone Genes ANEVA-DOT Genes in Variant and Haplotype Aggregated ASE Data. 2025. DOI: 10.5281/zenodo.15226389.

26. Ganapathy, K.R. and P. Mohammadi ANEVAh VG Estimates for GTEx and MAGE. 2025. DOI: 10.5281/zenodo.15226130.

27. Ganapathy, K.R. and P. Mohammadi ASE Data for MAGE. 2025. DOI: 10.5281/zenodo.15226193.

28. Pereira, L., et al., African genetic diversity and adaptation inform a precision medicine agenda. Nature Reviews Genetics, 2021. 22(5): p. 284–306.

29. Reed, F.A. and S.A. Tishkoff, African human diversity, origins and migrations. Current opinion in genetics & development, 2006. 16(6): p. 597–605.

30. Ashraf, B. and D.J. Lawson, Genetic drift from the out-of-Africa bottleneck leads to biased estimation of genetic architecture and selection. European Journal of Human Genetics, 2021. 29(10): p. 1549–1556.

31. Weir, B.S. and C.C. Cockerham, Estimating F-statistics for the analysis of population structure. evolution, 1984: p. 1358–1370.

32. Ding, Y., et al., Polygenic scoring accuracy varies across the genetic ancestry continuum. Nature, 2023. 618(7966): p. 774–781.

33. Collins, R.L., et al., A cross-disorder dosage sensitivity map of the human genome. Cell, 2022. 185(16): p. 3041–3055.e25.

34. Firth, H.V., et al., DECIPHER: database of chromosomal imbalance and phenotype in humans using ensembl resources. The American Journal of Human Genetics, 2009. 84(4): p. 524–533.

35. Virtanen, P., et al., SciPy 1.0: fundamental algorithms for scientific computing in Python. Nature methods, 2020. 17(3): p. 261–272.

36. Andrews, S., FastQC: A Quality Control Tool for High Throughput Sequence Data. 2010.

37. Zhao, H., et al., CrossMap: a versatile tool for coordinate conversion between genome assemblies. Bioinformatics, 2014. 30(7): p. 1006–1007.

38. Danecek, P., et al., Twelve years of SAMtools and BCFtools. Gigascience, 2021. 10(2): p. giab008.

39. Byrska-Bishop, M., et al., High-coverage whole-genome sequencing of the expanded 1000 Genomes Project cohort including 602 trios. Cell, 2022. 185(18): p. 3426–3440. e19.

40. Li, H., Tabix: fast retrieval of sequence features from generic TAB-delimited files. Bioinformatics, 2011. 27(5): p. 718–719.

41. Picard toolkit. 2019, Broad Institute.

42. Li, H. and R. Durbin, Fast and accurate long-read alignment with Burrows–Wheeler transform. Bioinformatics, 2010. 26(5): p. 589–595.

43. Di Tommaso, P., et al., Nextflow enables reproducible computational workflows. Nature biotechnology, 2017. 35(4): p. 316–319.

44. Dobin, A., et al., STAR: ultrafast universal RNA-seq aligner. Bioinformatics, 2013. 29(1): p. 15–21.

45. Rozowsky, J., et al., AlleleSeq: analysis of allele-specific expression and binding in a network framework. Molecular systems biology, 2011. 7(1): p. 522.

46. Li, B. and C.N. Dewey, RSEM: accurate transcript quantification from RNA-Seq data with or without a reference genome. BMC bioinformatics, 2011. 12: p. 1–16.

47. Köster, J. and S. Rahmann, Snakemake—a scalable bioinformatics workflow engine. Bioinformatics, 2012. 28(19): p. 2520–2522.

48. Trapnell, C., L. Pachter, and S.L. Salzberg, TopHat: discovering splice junctions with RNA-Seq. Bioinformatics, 2009. 25(9): p. 1105–1111.

49. Van De Geijn, B., et al., WASP: allele-specific software for robust molecular quantitative trait locus discovery. Nature methods, 2015. 12(11): p. 1061–1063.

50. Quinlan, A.R. and I.M. Hall, BEDTools: a flexible suite of utilities for comparing genomic features. Bioinformatics, 2010. 26(6): p. 841–842.

51. Zaidi, S., et al., De novo mutations in histone-modifying genes in congenital heart disease. Nature, 2013. 498(7453): p. 220–223.

52. Van der Auwera, G.A., et al., From FastQ data to high-confidence variant calls: the genome analysis toolkit best practices pipeline. Current protocols in bioinformatics, 2013. 43(1): p. 11.10. 1-11.10.33.

53. McKenna, A., et al., The Genome Analysis Toolkit: a MapReduce framework for analyzing next-generation DNA sequencing data. Genome research, 2010. 20(9): p. 1297–1303.

54. Chang, C.C., et al., Second-generation PLINK: rising to the challenge of larger and richer datasets. Gigascience, 2015. 4(1): p. s13742-015-0047-8.

55. Delaneau, O., et al., Accurate, scalable and integrative haplotype estimation. Nat. Commun. 10, 5436. 2019.

56. Fort, A., et al., MBV: a method to solve sample mislabeling and detect technical bias in large combined genotype and sequencing assay datasets. Bioinformatics, 2017. 33(12): p. 1895–1897.

57. Favalli, V., et al., Machine learning-based reclassification of germline variants of unknown significance: The RENOVO algorithm. The American Journal of Human Genetics, 2021. 108(4): p. 682–695.

58. Wang, K., M. Li, and H. Hakonarson, ANNOVAR: functional annotation of genetic variants from high-throughput sequencing data. Nucleic acids research, 2010. 38(16): p. e164–e164.

59. Ganapathy, K.R. and P. Mohammadi Synthetically Contaminated ASE Data for Benchmarking aseQC. 2025. DOI: 10.5281/zenodo.15226467.

60. Waskom, M.L., Seaborn: statistical data visualization. Journal of Open Source Software, 2021. 6(60): p. 3021.

61. Danecek, P., et al., 1000 Genomes Project Analysis Group. 2011. The variant call format and VCFtools. Bioinformatics, 2011. 27(15): p. 2156–2158.

