## Supplementary Materials for "Improved Identification of Large-effect Rare Genetic Variants using Haplotype Aggregated Allele-specific Expression Data"

### Deriving $V^G$ Estimates for Haplotype Aggregated ASE Data

Due to the natural biological variation within the human population, it is common for hundreds of genes to exhibit evidence of allelic imbalance in any given transcriptome. This variation is further compounded by the binomial variation introduced by the count-based nature of ASE data. To quantify the expected extra-binomial variation in a given gene in a population, we use a binomial-logit-normal distribution to model haplotype-aggregated population ASE data for each gene. Specifically, we model the haplotype-specific expression associated with the first haplotype (h1) observed for a gene in an individual, $eh1$, using a binomial distribution specified by total coverage n (that is $eh1+eh2$ for each gene), and the true, latent, allelic ratio, r associated with the first haplotype is:

$eh1 \sim$ Binomial$\mathrm{n, r}$ (1)

where the logit-transformed haplotype ratio, $sr$, is specified by a normal distribution:

$sr = \log2r\log21-r \sim$ Normal$s0, \sigma2$ (2)

where $s_0$ is the log2 allelic fold change associated with bias towards h1, and $\sigma 2$ is the standard deviation of the presumed genetic extra-binomial variation in ASE data in log2 allelic fold change scale. Note that in this model haplotype labels h1 and h2 are arbitrary in each individual, and thus, $s_0$ is expected to be zero. However, in practice, this assumption can be violated depending on the implementation of the phasing and haplotype aggregation tool used, such that the haplotype-level data for h1 may still have the tendency to represent the reference allele across a majority of individuals (or vice versa). Thus, we retain, the allelic bias term s 0 in the model to capture any reference bias confounding haplotype-aggregated data. We used gradient descent as implemented in optim package in R using the L-BFGS-B approach to maximize likelihood for parameters $s_0$ and $\sigma 2$ for each gene in a population:

$\mathrm{py} = \mathrm{yI}; I, s0, \sigma2 = i\mathrm{BinIi}; I, \mathrm{rLNr}; s0, \sigma2\mathrm{dr}$ (3)

The expected variance in allelic expression due to genetic variation in regulation is computed as

$VG = \ln esr+1-\mathrm{E}\ln e2psrdsr$ (4)

Where:

$E\ln e = \ln esr+1psrdsr$ (5)

and $psr =$ Normal$s0, \sigma2$. This approach offers a tractable and scalable framework for quantifying $V^G$ from haplotype-level data. To assess the uncertainty of the $V^G$ estimates under this simplified model, we compute parametric 95% bootstrap confidence intervals using the boot package in R [2]. Scripts for enabling $V^G$ estimation are available both within the provided container and as part of a standalone R package [3, 4].

### Supplementary Note on Clinical Cases

**True Positive Muscular Dystrophy Cases Uniquely Identified by ANEVA-h**

**Case D1**

D1, a patient with congenital muscular dystrophy, harbored a ~450 kb deletion in LARGE1, previously shown to result in aberrant splicing (Fig S9, [5]). Our previous variant-level ASE analysis, which captures the most highly expressed SNP within a gene, detected no significant allelic imbalance (26 reads to the reference, 19 to the alternate allele) [1]. However, haplotype-level analysis using ANEVA-h revealed complete monoallelic expression from the reference haplotype (21 reads) and none from the alternate (Table S3). This discrepancy likely reflects complexities in transcript processing and expression dynamics, where a single high-expression SNP does not fully represent allele-wide disruptions. By aggregating ASE across phased haplotypes, ANEVA-h identified a novel imbalance missed at the variant level data, highlighting its power in detecting hidden regulatory disruptions.

**Prioritized Gene/Variant Pairs among Undiagnosed Muscular Dystrophy Cases by ANEVA-h**

**Case N19**

N19, a patient with suspected congenital muscular dystrophy, remained undiagnosed following standard exome and RNA-sequencing analyses [1, 5]. Previous analysis using ANEVA and variant-level ASE revealed significant allelic imbalance in ENO3, a gene primarily involved in glycolytic metabolism [1, 6]. No rare or potentially pathogenic variants were identified in ENO3 using RENOVO, and the clinical relevance of this signal remains uncertain. Subsequent application of ANEVA-h using haplotype-level ASE identified significant signal in NEB, a gene with well-established connections to congenital myopathies (Table S3, [1, 5, 7, 8]). ASE analysis revealed 273 reads assigned to haplotype 1 and 613 reads to haplotype 2, indicating a marked allelic imbalance (Table S3). Despite the gene's phenotypic relevance, variant prioritization (via RENOVO) across coding, splicing, and annotated regulatory regions did not reveal any rare or high-confidence pathogenic variants. The absence of explanatory variation in the presence of strong allelic imbalance suggests that pathogenicity may be mediated by mechanisms not captured through standard short-read sequencing approaches, including deep intronic mutations, disruption of distal regulatory elements, epigenetic silencing, or cryptic structural variation [9]. Thus, the case has been subsequently flagged for follow-up, with NEB being prioritized for further investigation (Table S3).

**Variant-Level True Positives Not Captured in Haplotype-Level ANEVA-h**

**Case C11**

C11, a patient with a congenital fiber-type disproportion phenotype, harbored a pathogenic missense variant (rs145088074) and an exonic synonymous splicing variant (rs1234999215) in the causal gene RYR1 [1]. Our previous variant-level ASE analysis revealed significant allelic imbalance, with 1,362 reads (61.5%) mapping to the reference allele and 851 reads (38.4%) to the alternate allele at the missense SNP rs145088074, indicating markedly aberrant ASE from ANEVA-DOT (Table S3, [1]). In contrast, haplotype-level analysis using ANEVA-h aggregated ASE across multiple variants, yielding a total of 7,161 reads—3,184 (44.4%) from haplotype 1 and 3,987 (55.5%) from haplotype 2 —without evidence of aberrant ASE in ANEVA-DOT (Table S3). Notably, we observed that the causal ASE-SNP (rs145088074) lacked allelic data in RYR1 in the haplotype-level output. Further investigation revealed that the genotype had been lost while phasing through population-based methods, leading to the loss of significant ASE signal.

**Case D12**

D12, a patient with an Ehlers-Danlos syndrome Type VI phenotype, harbored a causal deleterious nonsense mutation (rs121913552) and a missense mutation (rs199730384) [1]. Our previous variant-level ASE analysis revealed significant allelic imbalance, with 10 reads (12.6%) mapping to the reference allele and 69 reads (87.3%) to the alternate allele at the nonsense variant, which exhibited the highest expression in the gene [1]. In contrast, haplotype-level ASE analysis using PAC aggregated ASE across multiple variants, yielding a total of 116 reads—50 (43.1%) from the haplotype 1 and 66 (56.8%) from haplotype 2 —without evidence of significant ASE imbalance (Table S3). Notably, we observed that the causal ASE-SNP (rs121913552) lacked allelic data in PLOD1 in the haplotype-level output. Further investigation revealed that the variant was not present in the population-phased genotypes for the sample. This suggests that the variant (and its associated ASE) had been once again lost while phasing due to limitations in the population-based phasing strategies.

**Prioritized Cases Among the Pediatric Congenital Heart Disease Cohort**

**Case 1-00596**

1-00596, is a newborn patient with congenital anomalies involving the craniofacial, cardiovascular, skeletal, and airway systems, including telecanthus, large eyes, a high-arched palate, a posterior neck nodule, structural heart abnormalities, and laryngeal anomalies. Diagnoses include complex congenital heart defects involving hypoplastic left heart syndrome and related abnormalities. Genetic analysis revealed a large run of homozygosity (ROH) on chromosome 2 but no chromosomal aneuploidy or 22q11 deletion. Haplotype-level ASE analysis demonstrated significant allelic imbalance in MYH7, with 133 reads (25.4%) arising from one haplotype and 391 reads (74.6%) from the other, as detected by ANEVA-DOT (Table S4). A rare missense variant, rs746724436 (chr14:23418358 C>T), exhibited detectable ASE and is absent or extremely rare in population databases (<0.01% in gnomAD, ExAC, TOPMed Bravo) [10, 11]. The variant has deleterious predictions from Revel and Cardioboost (CM) models and is classified as a variant of uncertain significance (VUS) in relation to hypertrophic and general cardiomyopathy in ClinVar ) [10, 12]. While MYH7 variants are frequently associated with cardiomyopathies, their role in congenital heart defects is less well established [13]. Parental genotyping confirmed heterozygosity in both parents, suggesting the observed ASE imbalance may result from compensatory expression favoring the common allele. The large ROH on chromosome 2 raises the possibility of additional mechanisms such as unmasked recessive variants, cis-regulatory effects, or imprinting disturbances [14]. Further investigation is thus warranted to determine the pathogenic role of this variant and other potential genetic contributors.

**Case 1-06936**

1-06396 is a 3-month-old patient with congenital cardiovascular anomalies, including a conotruncal abnormality and a ventricular septal defect. No additional abnormalities were noted in craniofacial, skeletal, airway, or systemic structures, and no dysmorphic facial features were observed. Genetic analysis identified a rare nonsense variant in MAP2K1 (chr15:66485037 G>A), absent from population databases such as gnomAD, ExAC, and TOPMed Bravo, and not indexed in ClinVar [10-12]. The variant has deleterious predictions from FATHMM-MKL, EIGEN PC, and BayesDel models [10, 11]. Haplotype-level ASE analysis revealed significant allelic imbalance in MAP2K1, with 47 reads (94%) arising from the reference haplotype and 3 reads (6%) from the alternate haplotype, indicating aberrant expression as detected by ANEVA-DOT (Table S4). MAP2K1 mutations are frequently associated with congenital heart defects and cardiofaciocutaneous syndrome, which typically includes characteristic craniofacial features, though such features were absent in this proband [15]. Parental genotyping confirmed inheritance of the variant from one parent, with genotype data missing for the other. The rarity of the variant, combined with its aberrant ASE and established links to MAP2K1-related phenotypes, suggests further functional and clinical evaluation is needed to assess its relevance to the proband’s presentation.

### Supplementary Images

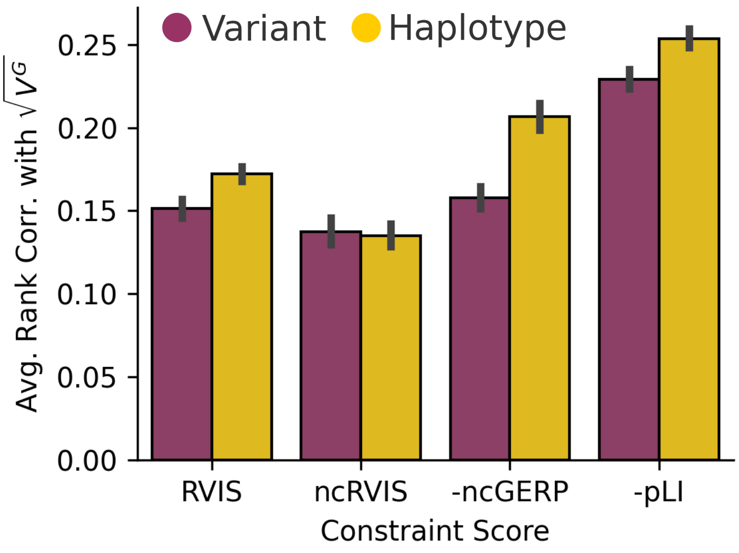


**Figure S1. Average Spearman Correlation Between $SD^G$ ($\sqrt{V^G}$) and Genomic Constraint Metrics**
Mean Spearman correlation of sample-level dosage variance (SDG, √VG) with coding constraint (RVIS, pLI), noncoding constraint (ncRVIS), and noncoding conservation (ncGERP), averaged across 49 GTEx tissues. Error bars represent the 95% confidence interval of the mean correlation across tissues.

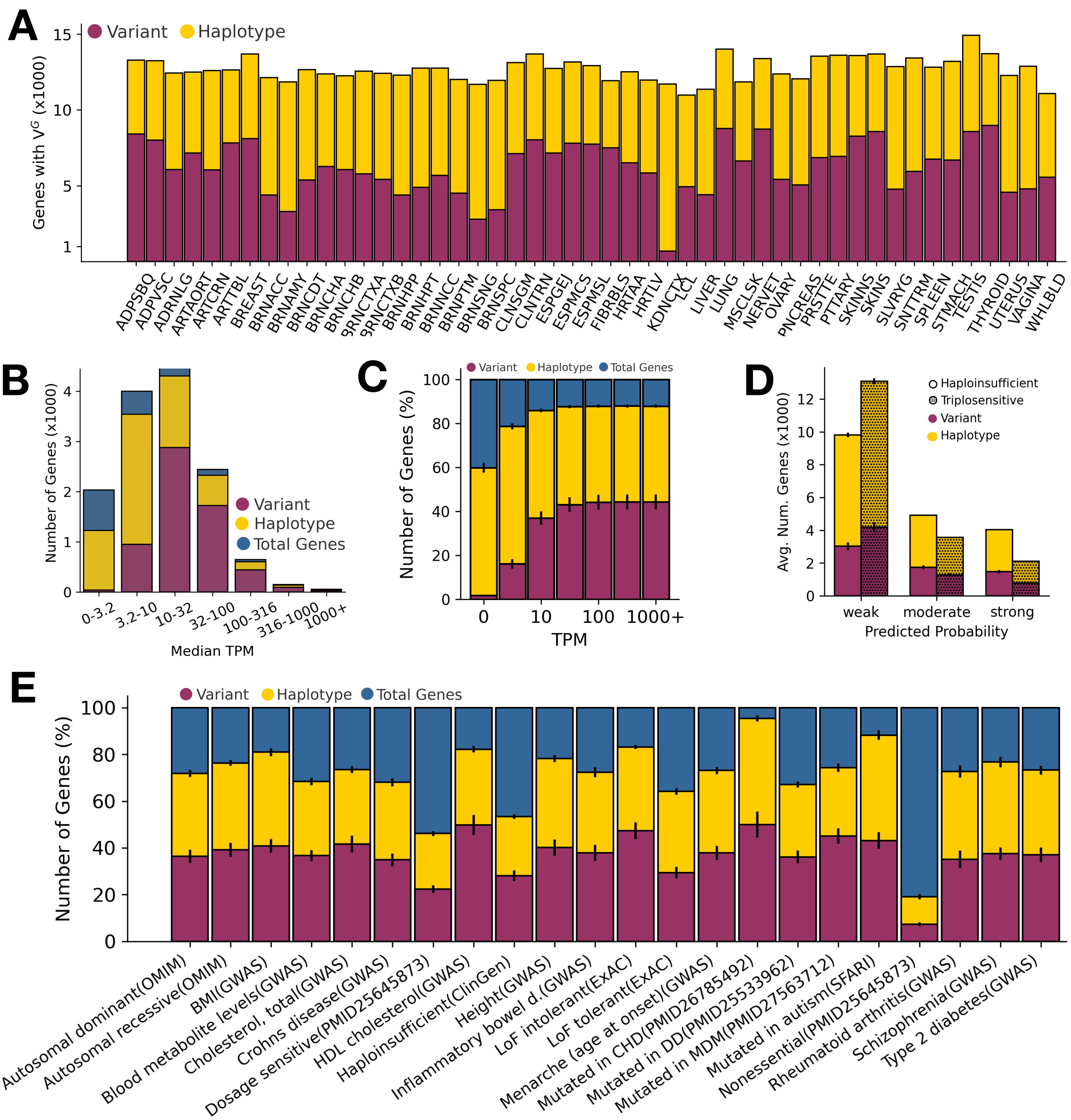


**Figure S2. Number of Genes with VG Estimates in Variant vs. Haplotype-Level ASE Data**

(A) Total number of protein-coding genes with VG estimates; (B) Number of genes with VG estimates across median TPM bins; (C) Cumulative proportion of genes with VG estimates as a function of TPM. Confidence Interval (CI) bars indicate 95% CI of the mean number of genes across 49 GTEx tissues; (D) Number of genes with VG estimates predicted to be haploinsufficient or triplosensitive. CI bars indicate 95% CI of the mean number of genes across 49 GTEx tissues; (E) Number of genes with VG estimates across different functional gene sets. CI bars indicate 95% CI of the mean number of genes across 49 GTEx tissues

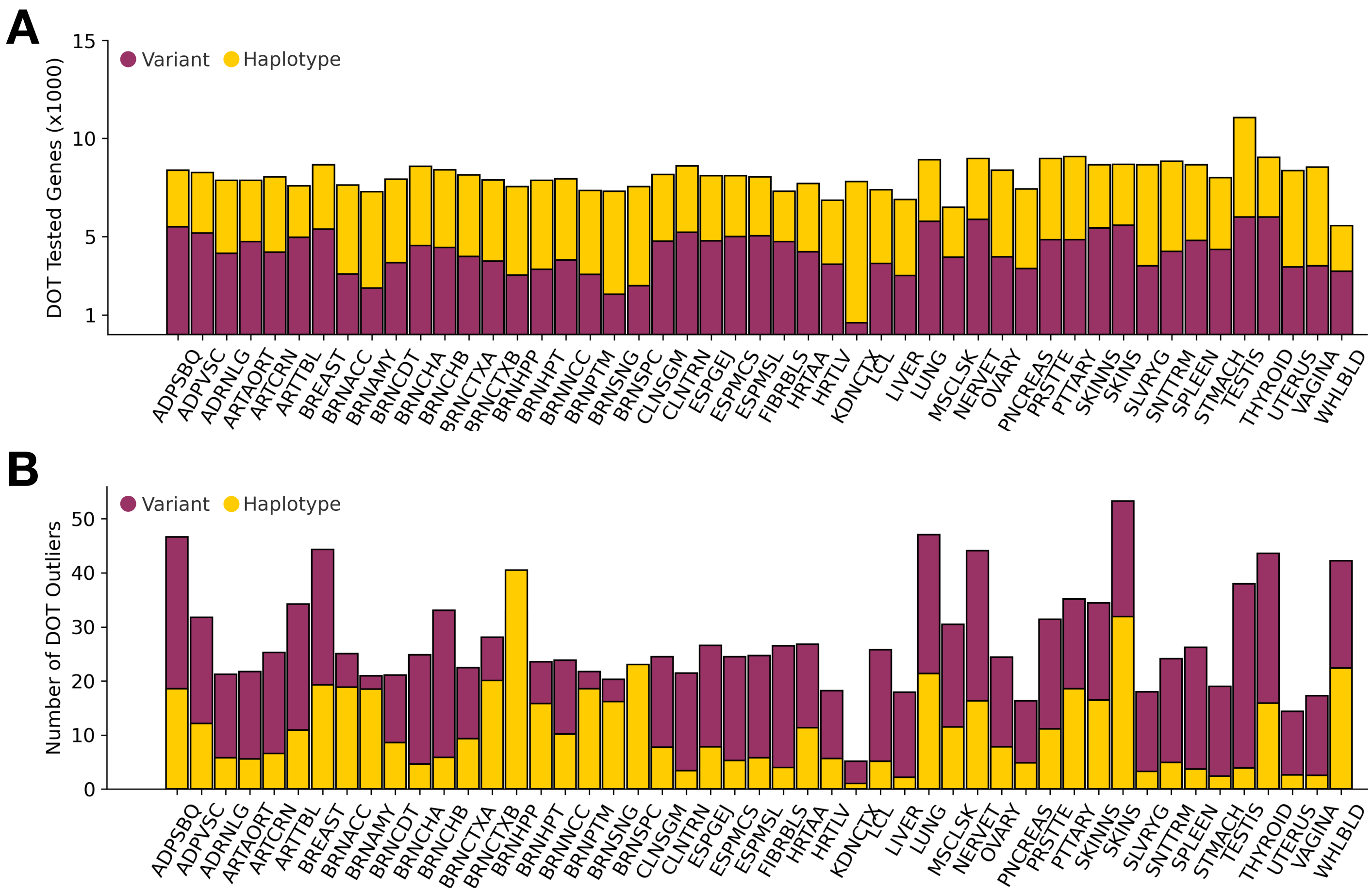


**Figure S3. Performance of ANEVA-DOT on Variant and Haplotype Level ASE Data**
(A) Average number of tested genes per sample across 49 GTEx tissues; (B) Average number of DOT outliers (FDR corrected; Benjamini-Hochberg method) per sample across 49 GTEx tissues.

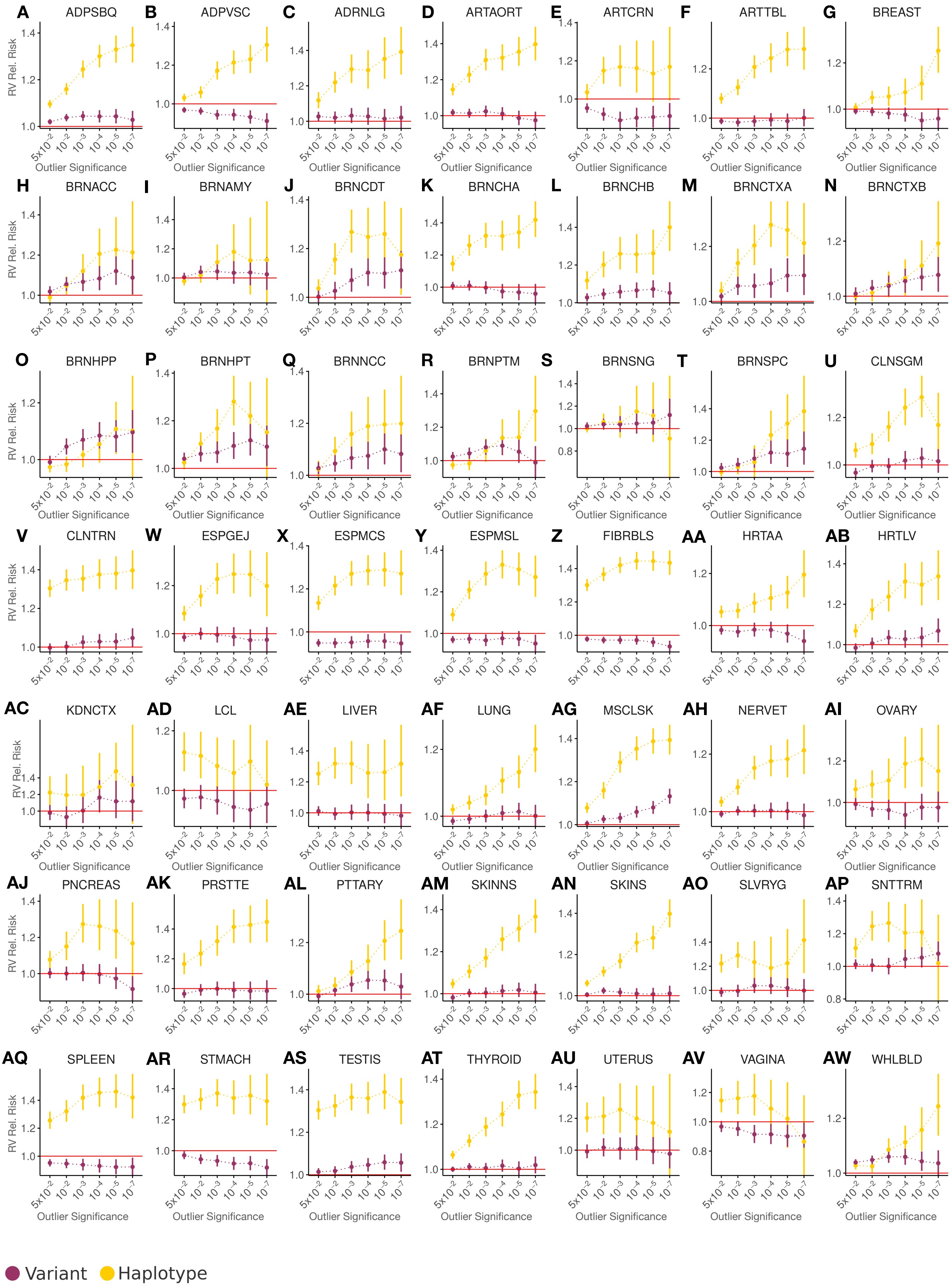
A ADPSBQ
B ADPVSC
C ADRNLG
D ARTAORT
E ARTCRN
F ARTTBL
G BREAST
H BRNACC
I BRNAMY
J BRNCDT
K BRNCHA
L BRNCHB
M BRNCTXA
N BRNCTXB
O BRNHPP
P BRNHPT
Q BRNNCC
R BRNPTM
S BRNSNG
T BRNSPC
U CLNSGM
V CLNTRN
W ESPGEJ
X ESPMCS
Y ESPMSL
Z FIBRBLS
AA HRTAA
AB HRTLV
AC KDNCTX
AD LCL
AE LIVER
AF LUNG
AG MSCLSK
AH NERVET
AI OVARY
AJ PNCREAS
AK PRSTTE
AL PTTARY
AM SKINNS
AN SKINS
AO SLVRYG
AP SNTTRM
AQ SPLEEN
AR STMACH
AS TESTIS
AT THYROID
AU UTERUS
AV VAGINA
AW WHLBLD
RV Rel. Risk
Outlier Significance
5x10^-2 10^-2 10^-3 10^-4 10^-5 10^-7
Variant
Haplotype

**Fig S4. Rare Variant Enrichment by Outlier Significance Before Removing False-positive Prone Genes** (A-AW) Comparison of changes to rare variant enrichment stratified by DOT outlier significance in variant- and haplotype-aggregated ASE data in 49 GTEx tissues.

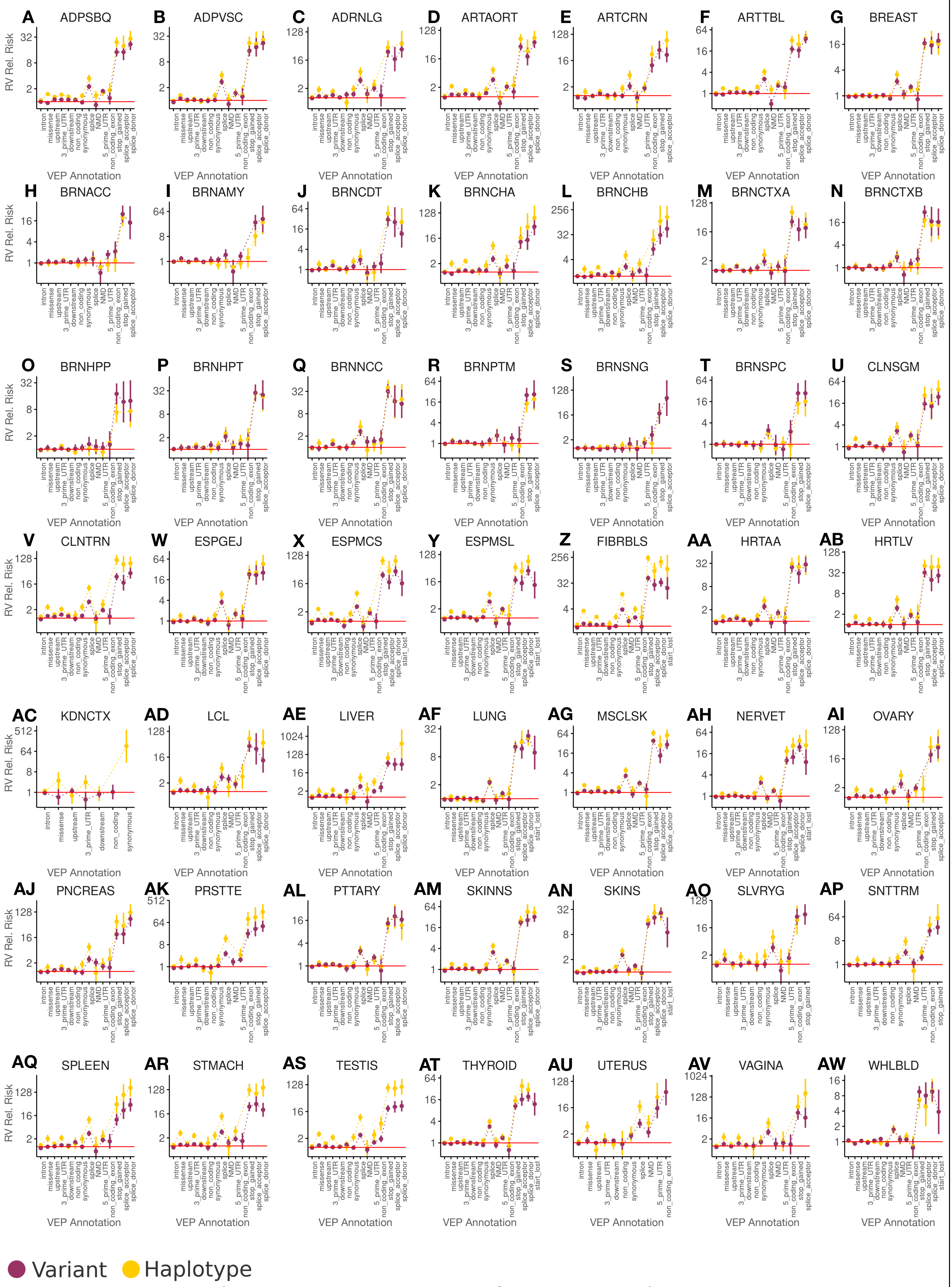


**Fig S5. Rare Variant Enrichment By VEP Consequence Before Removing False-positive Prone Genes**

(A-AW) Comparison of changes to rare variant enrichment among DOT outliers (FDR qval <= 0.05) stratified by VEP consequence in variant- and haplotype-aggregated ASE data in 49 GTEx tissues.

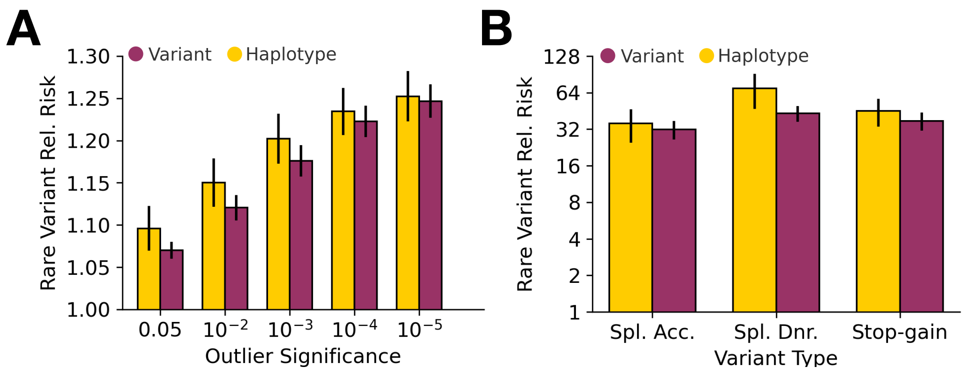


**Figure S6. Aggregated Rare Variant Enrichment Across GTEx Tissues After Removing False-Positive-Prone Genes**

(A) Aggregated relative risk of rare variant enrichment stratified by outlier significance (ANEVA-DOT p-value), with error bars representing the 95% confidence interval (CI) of the mean rare-variant relative risk across 49 GTEx tissues. (B) Aggregated relative risk of rare variant enrichment across high-impact VEP consequence categories, including only categories with available statistics in more than 45 GTEx tissues, with error bars representing the 95% CI of the mean rare-variant relative risk.

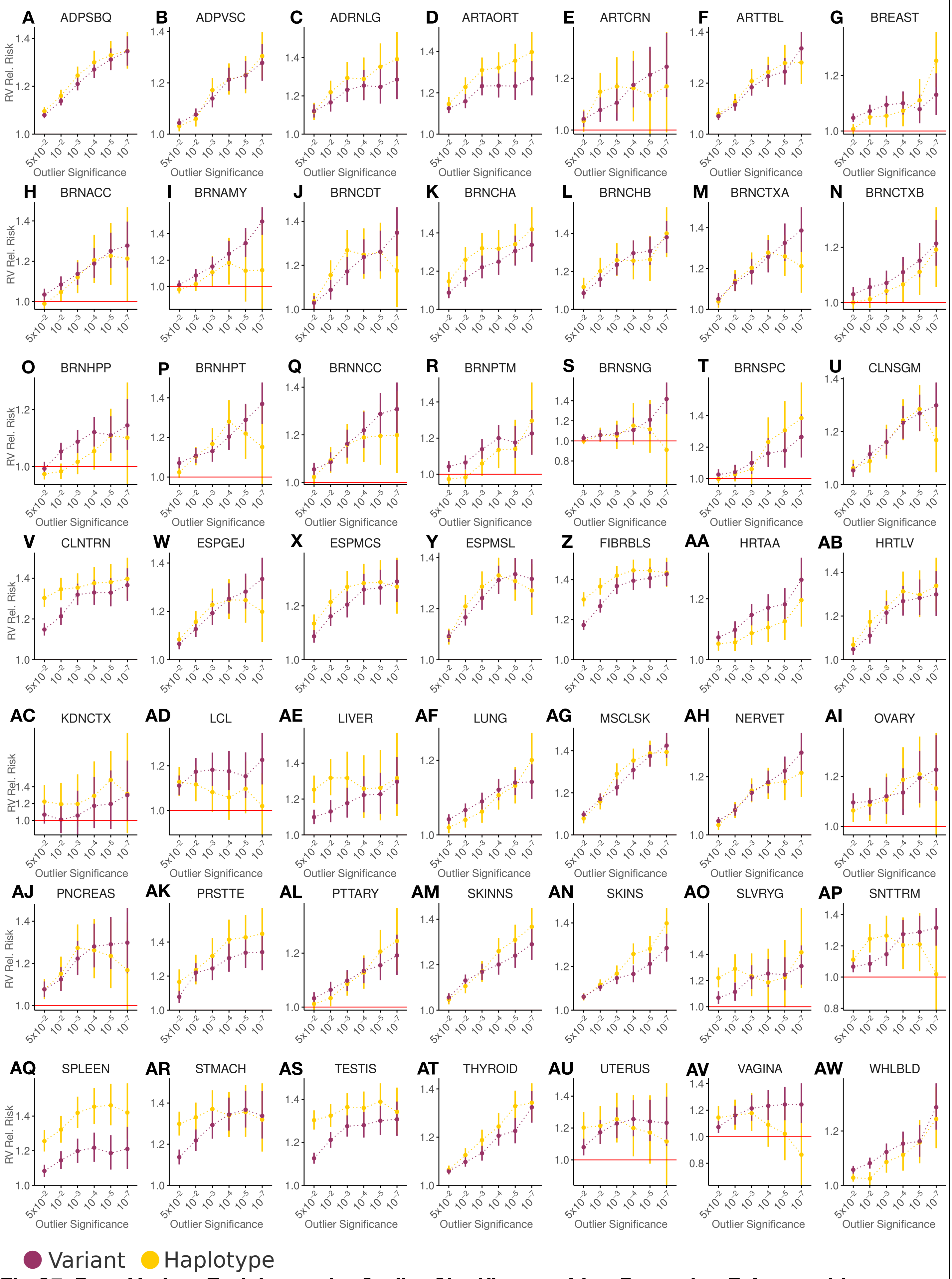


**Fig S7. Rare Variant Enrichment by Outlier Significance After Removing False-positive Prone Genes** (A-AW) Comparison of changes to rare variant enrichment stratified by DOT outlier significance in variant- and haplotype-aggregated ASE data in 49 GTEx tissues.

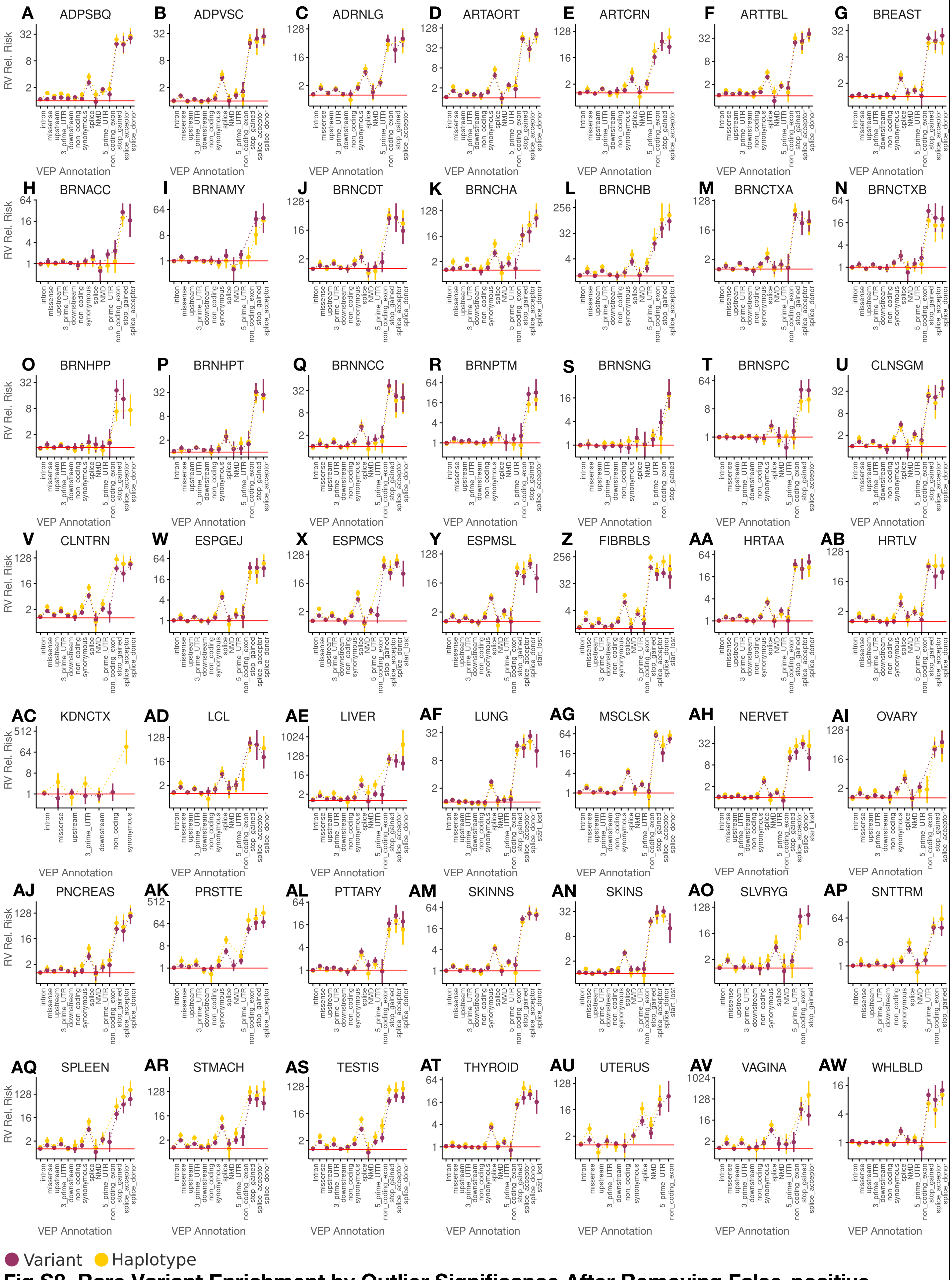


**Fig S8. Rare Variant Enrichment by Outlier Significance After Removing False-positive Prone Genes** (A-AW) Comparison of changes to rare variant enrichment among DOT outliers (FDR qval <= 0.05) stratified by VEP consequence in variant- and haplotype-aggregated ASE data in 49 GTEx tissues.

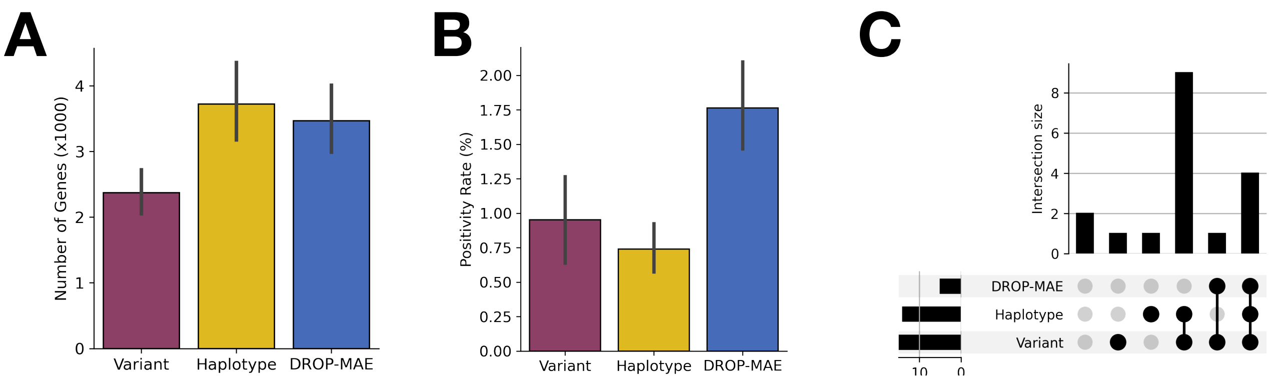


**Figure S9. Performance Assessment of ANEVA-h, ANEVA, and DROP-MAE in Positive Control Muscular Dystrophy Cases**

(A) Number of genes tested across ANEVA-h, ANEVA, and DROP-MAE, with error bars indicating the 95% confidence interval (CI) of the mean number of tested genes per case; (B) Test positivity rate, defined as the number of outlier genes per 100 tested genes, with error bars representing the 95% CI of the mean positivity rate across cases; (C) UpSet plot visualizing the number of positive control cases detected by each method and their overlap.

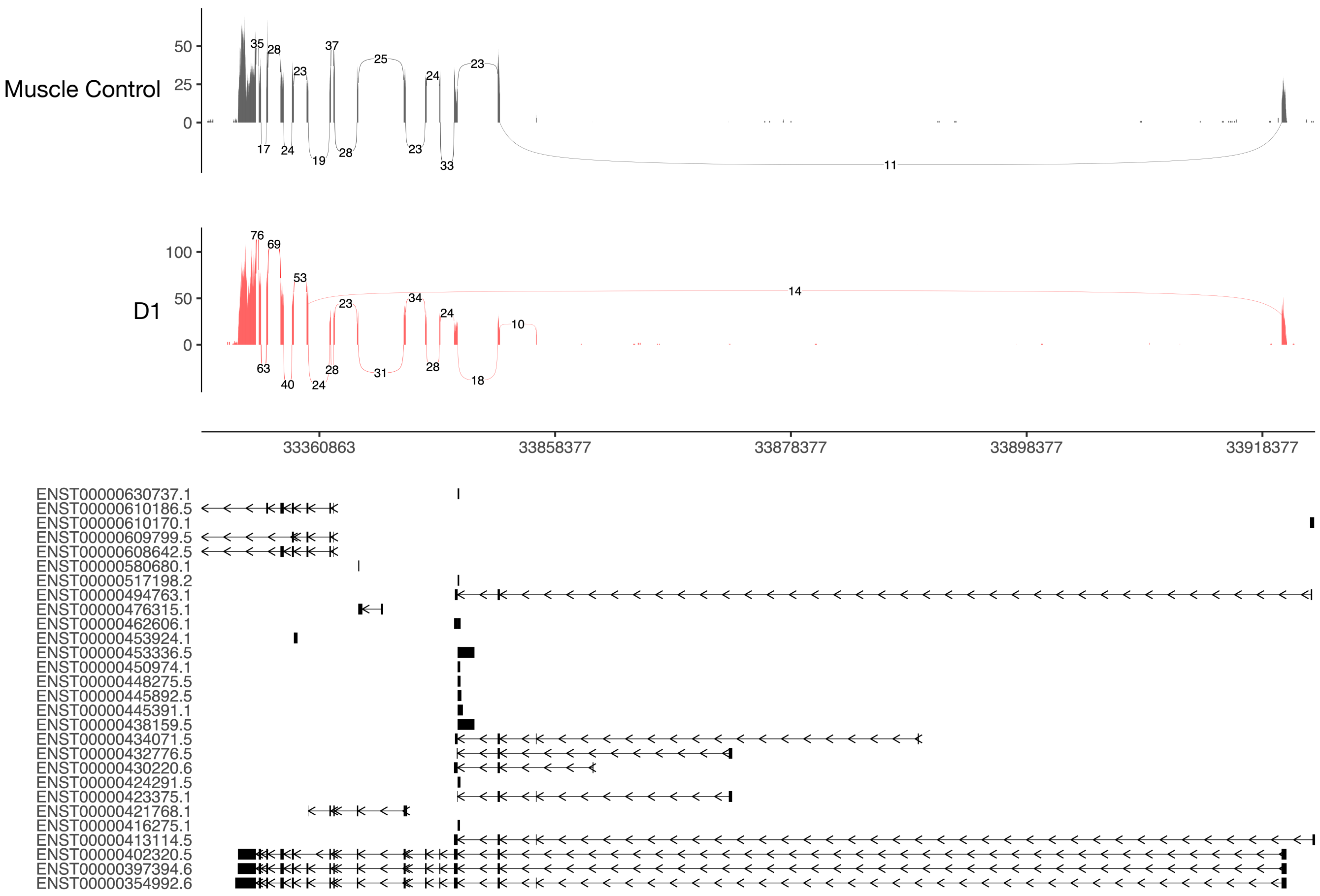


**Figure S10. Sashimi Plot of Causal Gene LARGE1 in Case D1**
Sashimi plot for the gene *LARGE1*, showing aberrant splicing due to a deletion in patient D1 (red) compared to a GTEx skeletal muscle control sample (black). Plots were generated using ggsashimi in R with GENCODE v26 annotations [16].

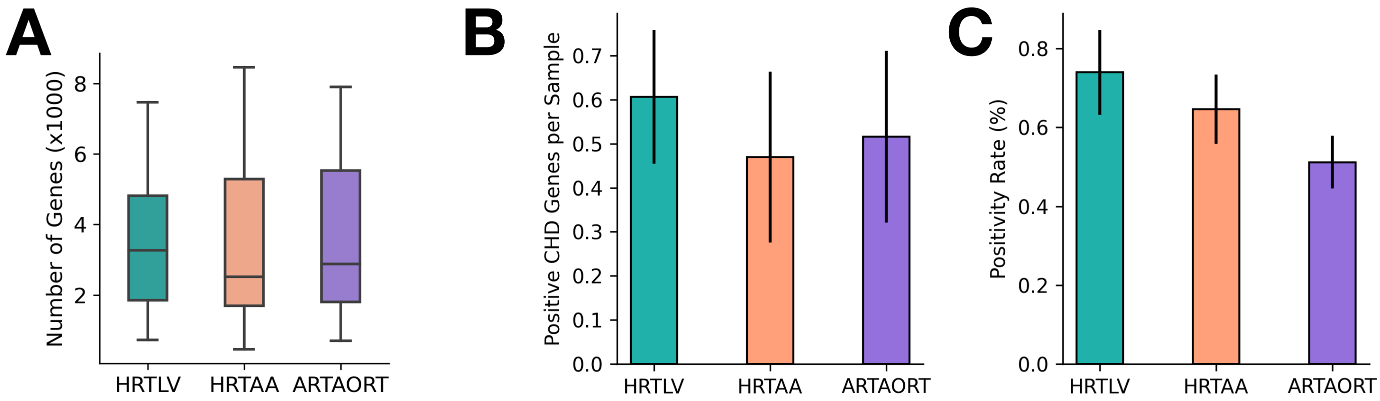


**Figure S11. Applying ANEVA-h to cases in PCGC cohort with haplotype ASE**
**(A)** Number of genes tested per sample stratified by GTEx tissue selection; **(B)** Positive CHD genes identified per sample; **(C)** Positivity rates for CHD cases.

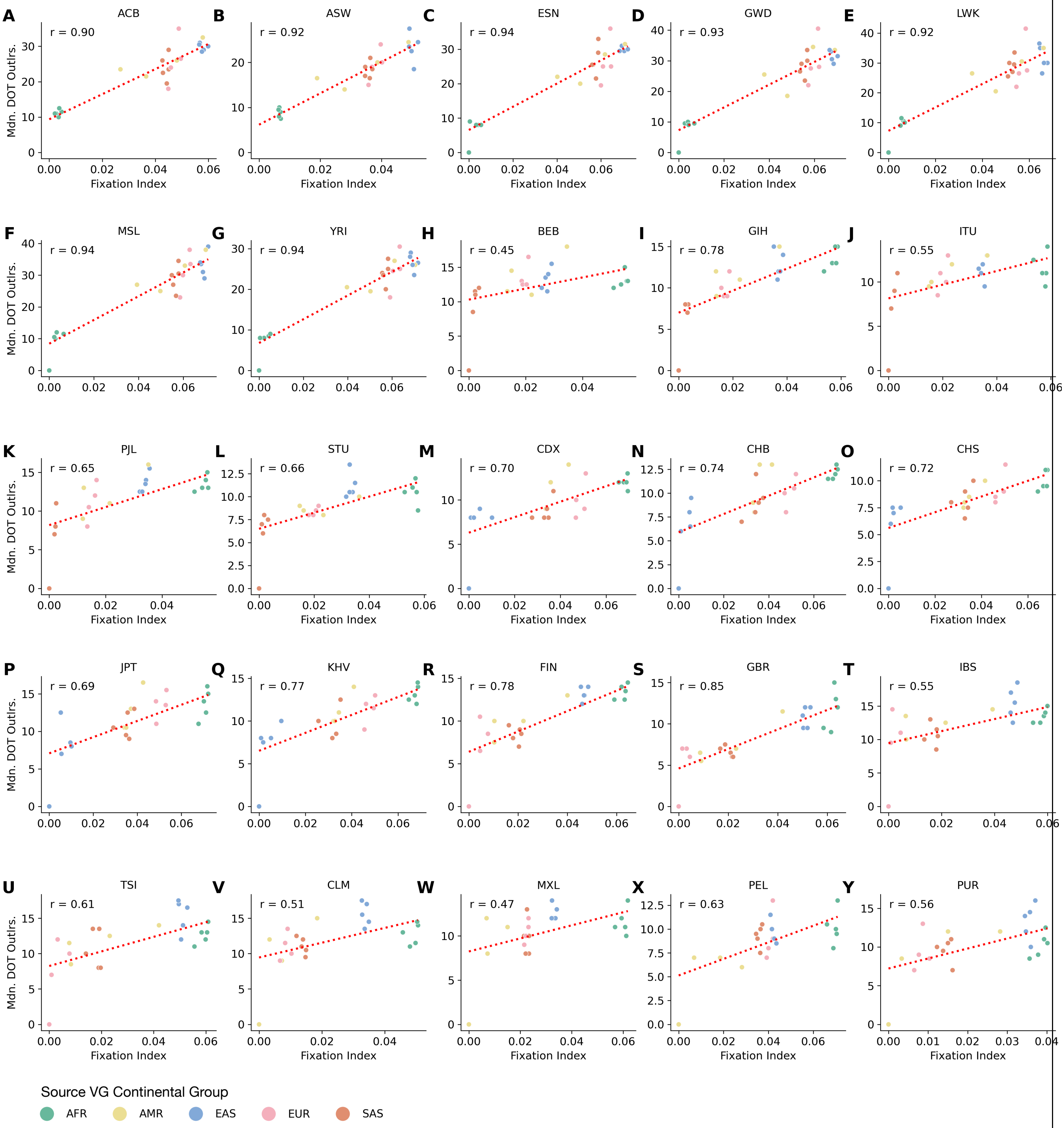


**Figure S12. Inflation of DOT Outliers with Increasing Genetic Distance from the Reference Population in the MAGE Dataset**

**(A-Y)** Median number of ANEVA-DOT outliers per sample across 25 distinct ancestry groups in MAGE , plotted as a function of the fixation index (Fst) of the source population, using VG estimates derived from diverse ancestral backgrounds.

### References


1. Mohammadi, P., et al., *Genetic regulatory variation in populations informs transcriptome analysis in rare disease.* Science, 2019. **366**(6463): p. 351-356.
2. Canty, A.J., *Resampling methods in R: the boot package.* The Newsletter of the R Project Volume, 2002. **2**(3): p. 2-7.
3. Song, E., P. Hoffman, and P. Mohammadi *ANEVA-h R Package*. 2025. DOI: 10.5281/zenodo.15226575.
4. Ganapathy, K.R. and P. Mohammadi *Singularity Container of Tools for Allele-Specific Expression Analysis*. 2025. DOI: 10.5281/zenodo.15226244.
5. Cummings, B.B., et al., *Improving genetic diagnosis in Mendelian disease with transcriptome sequencing.* Science translational medicine, 2017. **9**(386): p. eaal5209.
6. Hannah, W.B., et al., *Glycogen storage diseases.* Nature Reviews Disease Primers, 2023. **9**(1): p. 46.
7. Labeit, S., C.A. Ottenheijm, and H. Granzier, *Nebulin, a major player in muscle health and disease.* The FASEB journal, 2011. **25**(3): p. 822.
8. Yuen, M. and C.A. Ottenheijm, *Nebulin: big protein with big responsibilities.* Journal of Muscle Research and Cell Motility, 2020. **41**(1): p. 103-124.
9. Conesa, A., et al., *A survey of best practices for RNA-seq data analysis.* Genome biology, 2016. **17**: p. 1-19.
10. *Frankin by Genoox*. 2024 [cited 2024 September 15]; Available from: https://franklin.genoox.com/.
11. Kopanos, C., et al., *VarSome: the human genomic variant search engine.* Bioinformatics, 2019. **35**(11): p. 1978-1980.
12. Harrison, S.M., et al., *Using ClinVar as a Resource to Support Variant Interpretation.* Current Protocols in Human Genetics, 2016. **89**(1): p. 8.16.1-8.16.23.
13. Ritter, A., et al., *MYH7 variants cause complex congenital heart disease.* American Journal of Medical Genetics Part A, 2022. **188**(9): p. 2772-2776.
14. Szpiech, Z.A., et al., *Long runs of homozygosity are enriched for deleterious variation.* The American Journal of Human Genetics, 2013. **93**(1): p. 90-102.
15. Scorrano, G., et al., *The Cardiofaciocutaneous syndrome: from genetics to prognostic–therapeutic implications.* Genes, 2023. **14**(12): p. 2111.
16. Garrido-Martín, D., et al., *ggsashimi: Sashimi plot revised for browser-and annotation-independent splicing visualization.* PLoS computational biology, 2018. **14**(8): p. e1006360.
